# Bayesian uncertainty quantification to identify population level vaccine hesitancy behaviours

**DOI:** 10.1101/2022.12.13.22283297

**Authors:** David J. Warne, Abhishek Varghese, Alexander P. Browning, Mario M. Krell, Christopher Drovandi, Wenbiao Hu, Antonietta Mira, Kerrie Mengersen, Adrianne L. Jenner

## Abstract

When effective vaccines are available, vaccination programs are typically one of the best defences against the spread of an infectious disease. Unfortunately, vaccination rates may be suboptimal for a prolonged duration as a result of slow uptake of vaccines by the public. Key factors driving slow vaccination uptake can be a complex interaction of vaccine roll-out policies and logistics, and vaccine hesitancy behaviours potentially caused by an inflated sense of risk in adverse reactions in some populations or community complacency in communities that have not yet experienced a large outbreak. In the recent COVID-19 pandemic, public health responses around the world began to include vaccination programs from late 2020 to early 2021 with an aim of relaxing non-pharmaceutical interventions such as lockdowns and travel restrictions. For many jurisdictions there have been challenges in getting vaccination rates high enough to enable the relaxation of restrictions based on non-pharmaceutical interventions. A key concern during this time was vaccine hestitancy behaviours potentially caused by vaccine safety concerns fuelled by misinformation and community complacency in jurisdictions that had seen very low COVID-19 case numbers throughout 2020, such as Australia and New Zealand. We develop a novel stochastic epidemiological model of COVID-19 transmission that incorporates changes in population behaviour relating to responses based on non-pharmaceutical interventions and community vaccine uptake as functions of the reported COVID-19 cases, deaths, and vaccination rates. Through a simulation study, we develop a Bayesian analysis approach to demonstrate that different factors inhibiting the uptake of vaccines by the population can be isolated despite key model parameters being subject to substantial uncertainty. In particular, we are able to identify the presence of vaccine hesitancy in a population using reported case, death and vaccination count data alone. Furthermore, our approach provides insight as to whether the dominant concerns driving hesitancy are related to vaccine safety or complacency. While our simulation study is inspired by the COVID-19 pandemic, our tools and techniques are general and could be enable vaccination programs of various infectious diseases to be adapted rapidly in response to community behaviours moving forward into the future.

## 1 Introduction

By January 2021, two COVID-19 vaccines with greater than 90% efficacy to reduce symptomatic infection risk were approved in the EU (Polack et al., 2020; Baden et al., 2021). As these vaccines were rolled out globally, success of vaccination programs crucially depended on enough of the population accepting a vaccine (Lincoln et al., 2022) to reach heard immunity. Unsurprisingly, most countries were concerned with the effect of vaccine hesitancy on vaccine uptake (Schwarzinger et al., 2021).

Vaccine hesitancy is generally defined as a delay in acceptance or refusal of vaccines despite availability (Olivera Mesa et al., 2022). Vaccine hesitancy threatens to substantially limit the impact of vaccination as demonstrated by population survey studies found that a non-negligible proportion of the worlds population would not accept a vaccine if available (Gupta and Verma, 2022; Lincoln et al., 2022), for example, a survey study by Walsh et al. (2022) indicated the 23.3% of Irish and 26.3% of UK respondents were vaccine hesitant or resistant. Early studies even showed that COVID-19 vaccine hesitancy was increasing as in the lead-up to mass vaccination campaigns (Schwarzinger et al., 2021).

A unique feature of the COVID-19 pandemic has been the rapid response from the mathematical and statistical modelling community. This modelling had a substantial (though not always positive) impact on the public health responses globally (Ferguson et al., 2020; Hendy et al., 2021; Mouvoh et al., 2020; Plank et al., 2022; Price et al., 2020). Epidemiological models of various levels of complexity have been used to understand transmission dynamics of SARS-CoV-2 (Wodarz et al., 2021) including hospitalisation and fatality rates (Davies et al., 2020; Verity et al., 2020), forecast epidemic peaks and hospital utilisation (Birrell et al., 2021), infer asymptomatic or undocumented case numbers (Li et al., 2020; Flaxman et al., 2020; Wesner et al., 2021), inform the implementation of non-pharmaceutical interventions (NPIs) (Chinazzi et al.,, 2020; Mokhtari et al., 2021; Bouchnita et al., 2021a), and assess the efficacy and compliance of NPIs across jurisdictions efficacy (Flaxman et al., 2020; Warne et al., 2020; Wibbens et al., 2020). Mathematical modelling of within-host dynamics of SARS-CoV-2 (Bouchnita and Jebrane, 2020; Bouchnita et al., 2021b) and its interaction with the immune system (Fadai et al., 2021; Jenner et al., 2021) also provided many insights into potential treatments and therapeutics for COVID-19. When vaccines became available, epidemiological models including vaccination effects were employed to understand the effects of vaccination on slowing the pandemic (Volpert et al., 2021), inform vaccination programs (Aniţa et al., 2021; Hogan et al., 2021; Li et al., 2021; Nguyen et al., 2021), and investigate the effects of easing restrictions under a partially vaccination population (Olivares and Staffetti, 2021; Le et al., 2022).

In contrast, most of the modelling work investigating COVID-19 vaccine hesitancy have depended solely on statistical models or machine learning methods using survey data (Schwarzinger et al., 2021; Lincoln et al., 2022; Gupta and Verma, 2022; Bass et al., 2022; Walsh et al., 2022). Such approaches have been used to assess the willingness of individuals to get vaccinated (Lincoln et al., 2022) and account for effects of negative vaccine media related to mistrust and vaccine lethality (Schwarzinger et al., 2021; Gupta and Verma, 2022). Other survey based studies investigated the significance of different information sources on an individuals attitudes toward vaccination (Bass et al., 2022; Reno et al., 2021; Walsh et al., 2022). Using the vaccination strategy of Hogan et al. (2021), Olivera Mesa et al. (2022) investigated the potential impact of vaccine hesitancy on the control of the pandemic and the relaxation of NPIs using data on vaccine hesitancy from population surveys.

While these investigations analysing survey data are insightful in identifying predictors of vaccine hesitancy, there are limitations to the survey approach. In particular, survey-based approaches often rely on detailed data being available, thus limiting the applicability of the model to other countries or jurisdictions (Lindvall and Rönnerstrand, 2022). Furthermore survey cannot model the effect that disease dynamics has on vaccine hesitancy and *vice versa*. Unfortunately, there have been few mathematical modelling studies that consider this dynamic. Some examples explore the impacts of information on the behaviour related to vaccine hesitancy (Buonomo, 2020; Oduro et al., 2021) and strategies to counter mis-information (Buonomo et al., 2022; Lau et al., 2021; Sinclair et al., 2022).

Following work by Buonomo (2020), Warne et al. (2020) and Le et al. (2022), we develop mathematical modelling and statistical analysis tools to quantify vaccine hesitancy behaviour patterns using reported COVID-19 case numbers, deaths and vaccination numbers. The such tools can be more rapidly and readily deployed during a vaccination campaign that survey-based analyses. We emphasize that our approach is not designed to replace survey studies into vaccine hesitancy, but rather complement these studies. We incorporate the effect of a single vaccine into a stochastic epidemiological model that accounts for changes in behaviour related to transmission (i.e., related to NPIs) and vaccine uptake. Changes in population behaviour are driven by reported COVID-19 data that result in feedback loops. We demonstrate that a variety of realistic dynamics can be produced with our model under different scenarios for NPI strategies, vaccination roll-outs, and hesitancy behaviours. To obtain insight into vaccine hesitancy behaviours in a real vaccination campaign, the model would be calibrated using report case data. However, a key concern for the use of modelling to understand epidemics is whether data are sufficiently informative to ensure key model parameters are identifiable Kretzschmar et al. (2022); Browning et al. (2020). In this work, we address this important and often overlooked aspect of mathematical modelling by applying Bayesian uncertainty quantification. We demonstrate through a simulation study that different hesitancy behaviours, such as complacency and vaccine safety concerns can be distinguished from each other using reported case data and vaccination counts. This framework has the potential to assist public health policy makers in identifying the dominating causes of vaccine hesitancy rapidly to complement results of survey data analysis (Lindvall and Rönnerstrand, 2022).

## 2 Materials and Methods

In this section, we present a framework to analyse vaccine hesitancy behaviour patterns using publicly available COVID-19 case data and vaccination counts. Our approach consists of a novel stochastic epidemiological model that accounts for changes in interaction behaviour driven by NPIs and changes in vaccination uptake based on hesitancy. Structural and practical identifiability analysis is applied to our model to investigate the potential for these data to better understand vaccine hesitancy behaviours.

### 2.1 Data Sources and Structure

Here we highlight some important features of publicly available reported case data that we consider in this work.

#### 2.1.1 Data Sources

While our work is not specific to the COVID-19 pandemic, there were several important data repositories that became available during this period that form a strong motivation our analysis. In particular, the COVID-19 case data from Johns Hopkins University Coronavirus Resource Centre (https://coronavirus.jhu.edu/), the European Centre for Disease Prevention and Control (https://www.ecdc.europa.eu/en/covid-19), and the Our World in Data project (https://ourworldindata.org/coronavirus), provided invaluable examples that we consider as exemplars for our work.

#### 2.1.2 Data Structure

For each of the the data sources provided above, the data at least contain a time-series of cumulative reported cases, deaths and vaccinations doses split by first dose and second dose. These counts are also split over different jurisdictions. There are a number of challenges it using this data to model the epidemic evolution and understanding hesitancy. Firstly, the cumulative confirmed cases do not represent the actual number of infections that have occurred in the population. Primarily this will be due to undetected asymptomatic cases, but could also be impacted by false positive or false negative test results. Second, the cumulative case recoveries needs to be estimated as a proportion of cumulative cases that did not die and are not currently active^1^. Finally, and most importantly for th study of vaccine hesitance, the cumulative case and death data is not split by vaccination status, therefore the distribution of vaccination status across cases and deaths also needs to be inferred.

Since this work is designed to evaluate the potential of our methods to identify hesitancy behaviour patterns, we rely on simulated data using a model with known hesitancy patterns. However, our modelling and simulated data processes are constructed in such a way that they are consistent with real epidemiological data repositories such as Johns Hopkins University (Johns Hopkins University, 2020) https://github.com/CSSEGISandData/COVID-19 or Our World in Data (Mathieu et al., 2020) https://github.com/owid. For our simulation study, we have selected key population and epidemiological parameters such that behaviour is qualitatively similar to the United Kingdom COVID-19 situation between 1st September 2020 and 2nd December 2020, however, our various hesitancy scenarios are all hypothetical and not conclusions are specifically drawn from case data itself.

#### 2.1.3 Ethics

Ethics approval was not required for this study as it is a simulation based study. Furthermore, the target data of our methodology are publicly available reported case, death and vaccination count data that do not contain any confidential or identifiable patient data.

### 2.2 Model Development

Here, we describe our epidemiological model that incorporates both the effects on NPIs and the effect of vaccination hesitancy. Our approach builds upon the model employed by Warne et al. (2020) and Le et al. (2022) as described in Section 2.2.1. We then extend this model to include the effects of vaccination and vaccine hesitancy behaviours in Section 2.2.2. Nomenclature is provided in Tables 1 and 2 for reference throughout theses sections. For full mathematical details of the models see the Supplementary Materials.

**Table 1:**
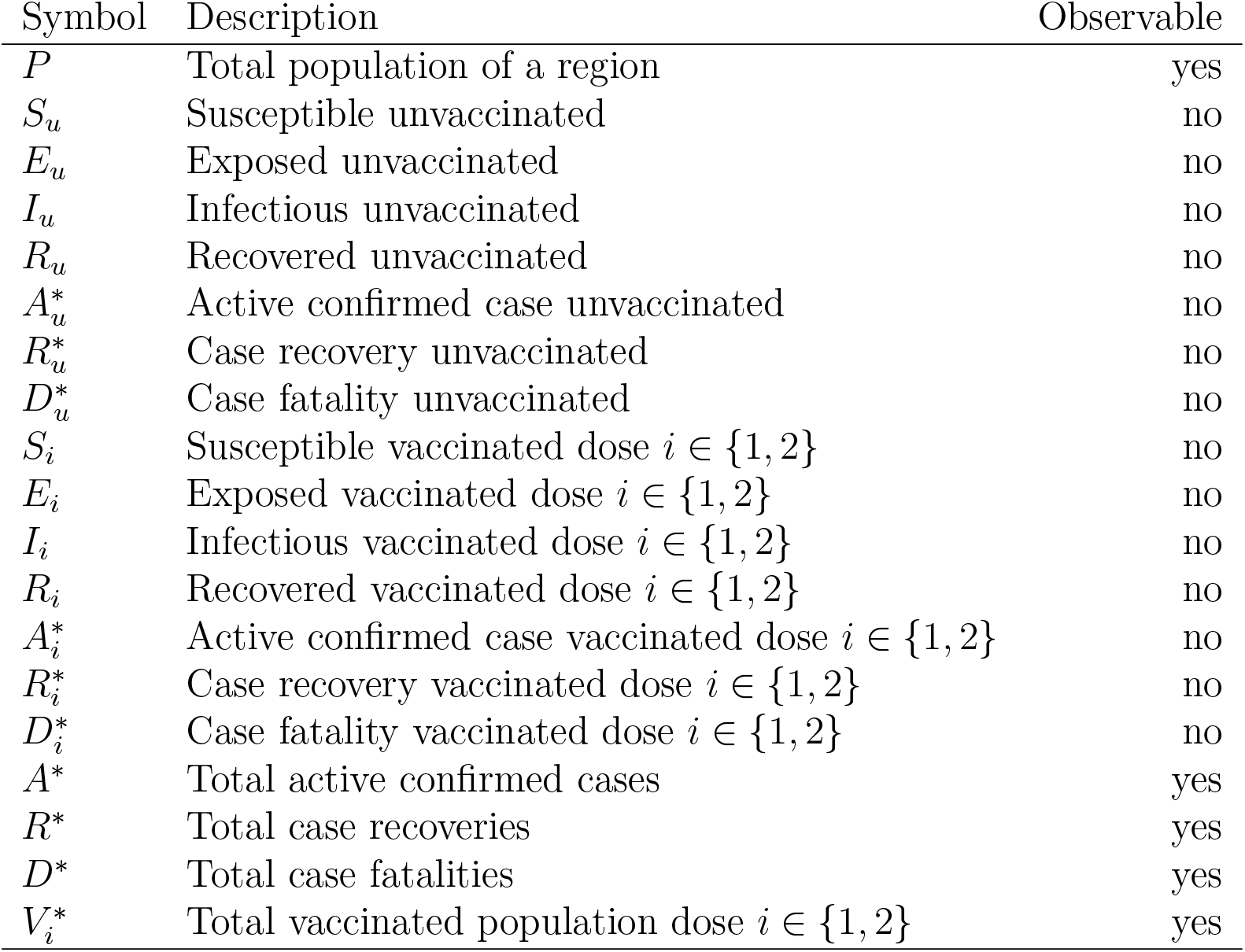
List of compartments in the full SEIR model including the case reporting process a two-dose vaccination program. A state is considered observable if it is available in a publicly available repository. States labelled with an asterisk “*” indicate data used in reporting, but aggregated before public release.

**Table 2:**
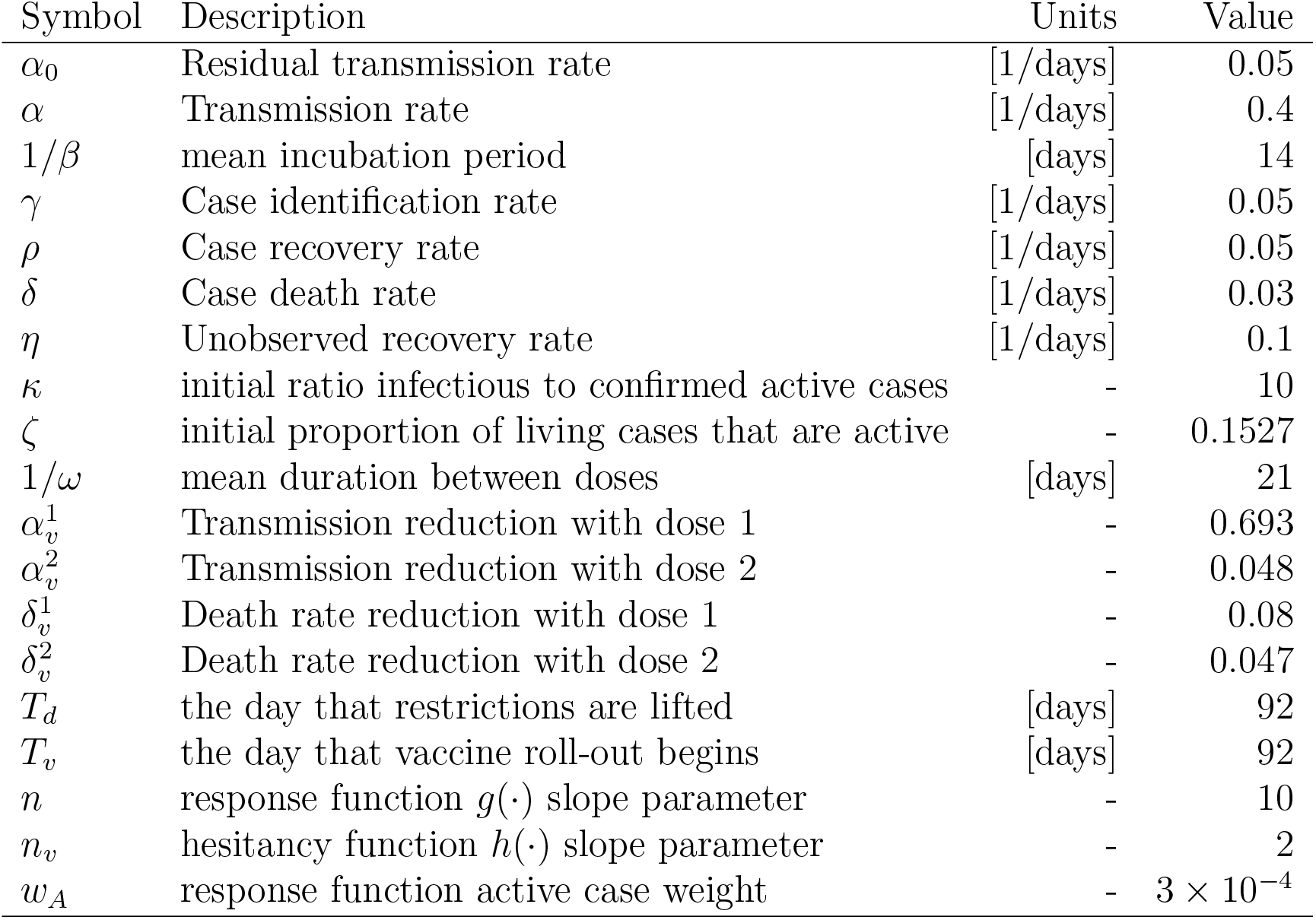
Model parameters fixed for synthetic dataset generation. Parameters related to vaccination rates and hesitancy are estimated via Bayesian inference. Parameter values are obtained from Warne et al. (2020) and Lopez Bernal et al. (2021b).

| Symbol | Description | Units | Value |
| --- | --- | --- | --- |
| $\alpha_0$ | Residual transmission rate | [1/days] | 0.05 |
| $\alpha$ | Transmission rate | [1/days] | 0.4 |
| $1/\beta$ | mean incubation period | [days] | 14 |
| $\gamma$ | Case identification rate | [1/days] | 0.05 |
| $\rho$ | Case recovery rate | [1/days] | 0.05 |
| $\delta$ | Case death rate | [1/days] | 0.03 |
| $\eta$ | Unobserved recovery rate | [1/days] | 0.1 |
| $\kappa$ | initial ratio infectious to confirmed active cases | - | 10 |
| $\zeta$ | initial proportion of living cases that are active | - | 0.1527 |
| $1/\omega$ | mean duration between doses | [days] | 21 |
| $\alpha_v^1$ | Transmission reduction with dose 1 | - | 0.693 |
| $\alpha_v^2$ | Transmission reduction with dose 2 | - | 0.048 |
| $\delta_v^1$ | Death rate reduction with dose 1 | - | 0.08 |
| $\delta_v^2$ | Death rate reduction with dose 2 | - | 0.047 |
| $T_d$ | the day that restrictions are lifted | [days] | 92 |
| $T_v$ | the day that vaccine roll-out begins | [days] | 92 |
| $n$ | response function $g(\cdot)$ slope parameter | - | 10 |
| $n_v$ | hesitancy function $h(\cdot)$ slope parameter | - | 2 |
| $w_A$ | response function active case weight | - | $3 \times 10^{-4}$ |

#### 2.2.1 Base epidemiological model

We consider a stochastic susceptible-exposed-infectious-recovered (SEIR) compartmental model based upon the work of Warne et al. (2020) and Le et al. (2022). In this framework, a population of size *P* is divided into four main epidemiological compartments: susceptible (*S*), exposed (*E*), infectious (*I*) and recovered (*R*). Since we cannot observe exactly the true number of active infectious individuals, these four components are considered unobservable or latent states. Infectious individuals will be identified as active confirmed cases (*A*\*) at rate *γ* > 0. These active confirmed cases will transition to reported deaths (*D*\*) or recoveries (*R*\*) at rates *δ* > 0 and *ρ* > 0 respectively. These three compartments are observable and correspond to reported COVID-19 data available from online dashboards and repositories such as Johns Hopkins University (Johns Hopkins University, 2020) https://coronavirus.jhu.edu/ or Our World in Data (Mathieu et al., 2020) https://ourworldindata.org/coronavirus. It should be noted that most data sources only reliably record the cumulative case numbers, *C*\* = *A*\* + *R*\* + *D*\*, and reported deaths, *D*\*. In addition, the reported *C*\* will be an underestimate of the true number of cumulative cases since infectious individuals can also recover without being reported as a confirmed case. In our model this transition occurs with rate *η* > 0.

In the SEIR model of the latent compartments, exposed individuals *E* will become infectious at rate *β* > 0 and those that are infectious can recover at rate *η* > 0. The population *R* represents cases in which the virus has run its course and the individual has received immunity without contributing to the COVID-19 case numbers; such is the case with asymptomatic cases that did not get tested. We do not explicitly model the possibility of a unreported death due to COVID-19, rather we absorbed this into the *R* compartment, that is, *R* is a removed category.The novelty of the approach by Warne et al. (2020) and Le et al. (2022) is the transmission mechanism in which a susceptible individual will be exposed with hazard rate [*α*_0_ + *αg*(·))]*I/P*, where *α*_0_ > 0 is the residual transmission rate, *α* > 0 is the transmission rate parameter that can be affected by changes in interaction behaviours of the population. These changes are modelled using a response function, *g*(·), that depends on the observables *A*\*, *D*\*, *R*\* and on time *t*. The feedback loop induced by the response function models the way the population changes behaviour in response external information such as public health advice, NPIs, or media reports. Similar approaches have also been considered for other diseases, such as influenza and HIV (Collinson and Heffernan, 2014; Teng et al., 2019). Figure 1 shows a schematic of the base model including the latent variables, *S, E, I* and *R*, and the feedback mechanism through *g*(·).

**Figure 1.**
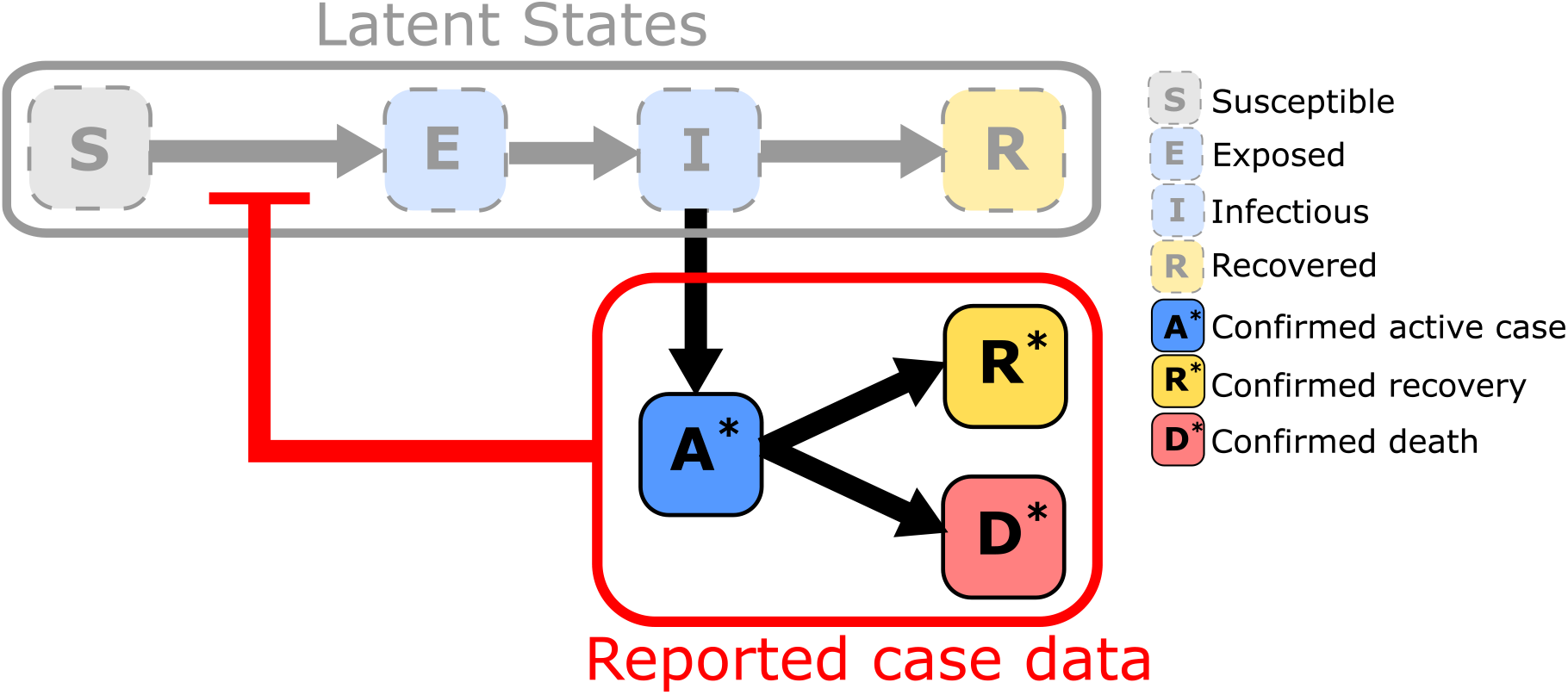
Schematic of the base epidemiological model. States are marked as labelled boxes and arrows indicate state transitions. The model consists of an unobservable (latent) SEIR model with the reported case data arising through the transition from *I* to *A*\*. The transmission is inhibited by a response function that depends on the observed reported case data.

Let 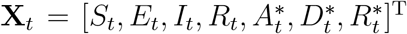 denote the state of the population at time *t* ∈ (0, *T*] with *t* = 0 at the initial condition and *t* = *T* at the last day of the time-series. Similarly, 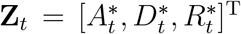 refers to the vector of observables. Under the assumption of a spatially homogeneous population and exponential waiting time between events, then the model is a discrete-state Markov process. Such a system can be simulated exactly using the Gillespie direct method or approximately using a time discretisation such as the *τ*-leaping scheme (Gillespie, 2001). In this work all stochastic simulations are performed using a *τ*-leaping scheme with *τ* = 1 day which is sufficiently accurate for our modelling purposes (Warne et al., 2020; Le et al., 2022).

To initialise simulations, typically only 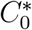 and 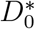 are available from data, we therefore denote two additional parameters *κ* ≥ 0 and *ζ* ∈ [0, 1] to initialise the other compartments. *κ* > 0 denotes the number of latent infections per active confirmed case and *ζ* is the proportion of alive cases that are currently active. Thus we arrive at initial states, 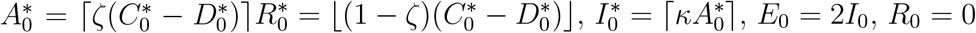, and 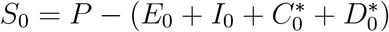.

Many different forms could be considered to describe different restriction strategies based on NPIs. For example in Warne et al. (2020), a lockdown strategy based on trigger threshold is implemented with

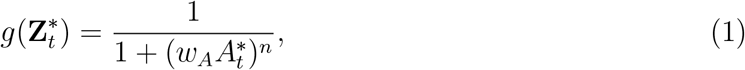

where *w*_*A*_ > 0 is a weight that affects the point in which the lockdown reaches 50% efficacy (that is *g*(1*/w*_*A*_) = 1*/*2) and *n* > 0 controls the rate at which the lockdown is introduced or eased (Figure 2(E)). That is, a larger weight corresponds to a rapid response (Figure 2(E) blue and red curves) and a smaller weight to a delayed response (Figure 2(E) yellow and purple curves). The slop parameter *n* > 0 relate to how sudden the introduction (resp. relaxation) of the response with larger *n* being a greater rate of introduction (Compare the yellow curve, *n* = 10, with the purple curve, *n* = 3, in Figure 2(E)). The choices for *w*_*A*_ and *n* have a substantial impact on the dynamics of the outbreak as shown in Figure 2(A)–(D). Figure 2 represents hypothetical scenarios initialised using confirmed case and death data for the United Kingdom on 1st September 2020 with 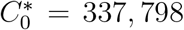 and 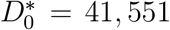. Unsurprisingly, we observe an early response leads to fewer cases and deaths than a delayed response (Compare Figure 2(A) with Figure 2(C) and Figure 2(B) with Figure 2 (D)). A sudden response leads to more frequent, higher peak, short duration epidemic waves, whereas a gradual response leads to infrequent, lower peak, long duration waves (Compare Figure 2(A) with Figure 2(B) and Figure 2(C) with Figure 2 (D)).

**Figure 2.**
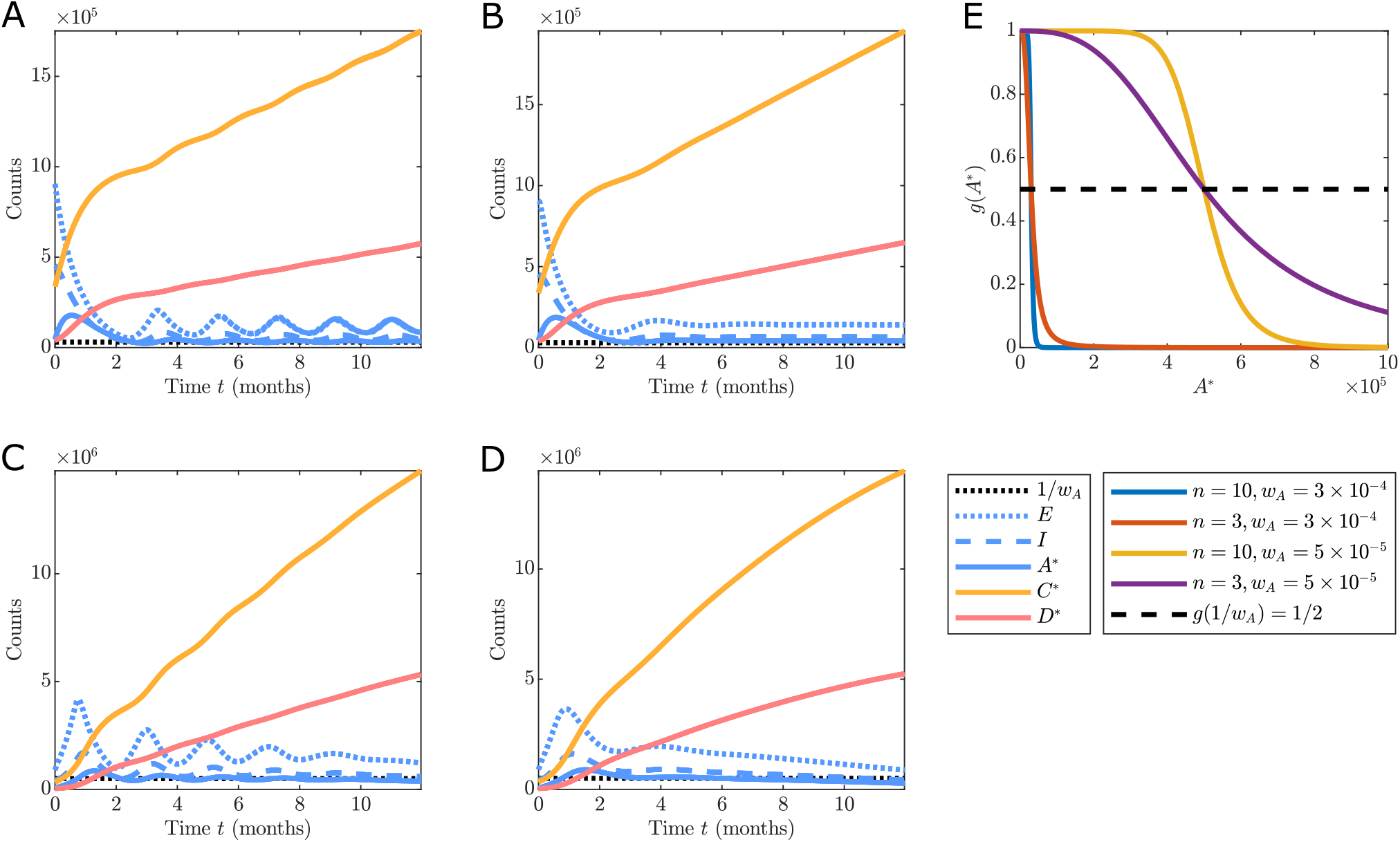
Realisations of the base epidemiological model for different configurations of the response function *g*(*A*\*) = 1/(1 + (*w*_*A*_*A*\*)^*n*^) with: (A) *n* = 10 and *w*_*A*_ = 3 *×* 10^−4^; (B) *n* = 3 and *w*_*A*_ = 3 *×* 10^−4^; (C) *n* = 10 and *w*_*A*_ = 5 *×* 10^−5^; and (D) *n* = 3 and *w*_*A*_ = 5 *×* 10^−5^. The shape of the response function in each case is shown in (E). Simulations are initialised based on the UK data on 1st September 2020 with 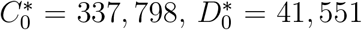 and model parameters are *α*_0_ = 0.05 [days^−1^], *α* = 0.4 [days^−1^], *β* = 0.07 [days^−1^], *γ* = 0.05 [days^−1^], *δ* = 0.03 [days^−1^], *ρ* = 0.05, *η* = 0.1 [days^−1^], *κ* = 10, and *ζ* = 0.15. The *τ*-leaping approximate stochastic simulation algorithm is used with time-step *τ* = 1 [days].

Multiphase behaviour in the response function can be introduced to simulate changes in intervention strategy over time. To do this we allow the response function to depend on time as well as on the observables. For example, consider a lockdown strategy following Equation (1) that is enacted over the interval *t* ∈ [0, *T*_*d*_], before easing all restrictions for *t > T*_*d*_. This could be implemented with the response function,

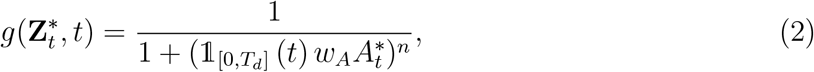

where 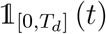 denotes an indicator function with 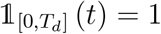 for *t* ∈ [0, *T*_*d*_] and 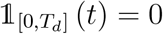 otherwise. In Figure 3(A)–(D), we use the same initial conditions as in Figure 2 and *T*_*d*_ = 92 corresponds to the 2nd of December 2020 when the second UK lockdown ended. As expected, we observe that regardless of the response applied for *t* ≤ *T*_*d*_, in the absence of vaccination, complete removal of restrictions for *t* > *T*_*d*_ leads to the same cumulative case and death counts.

**Figure 3.**
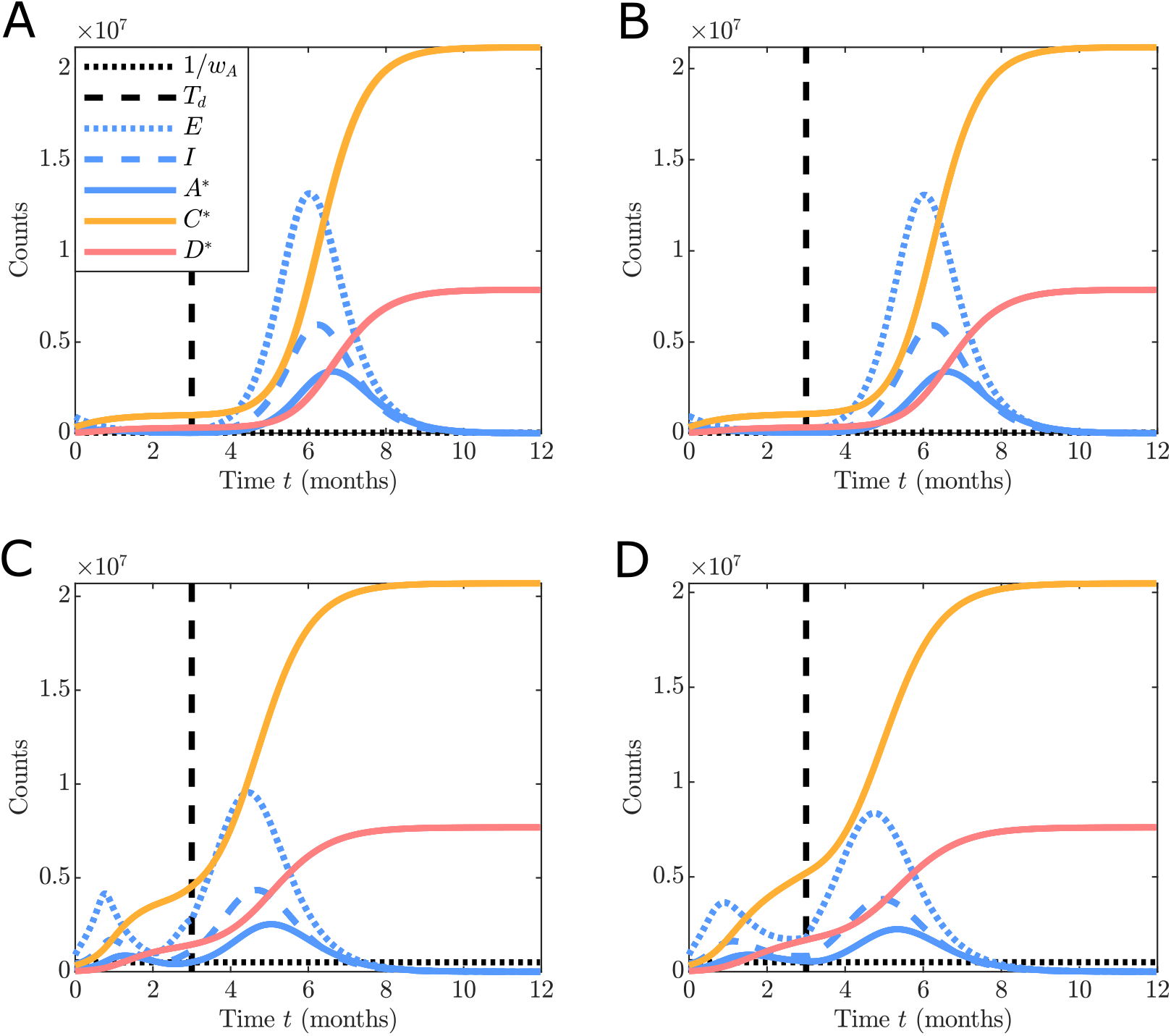
Realisations of the base epidemiological model for different configurations of the multiphase response function 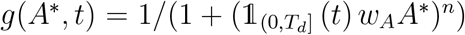 with: (A) *n* = 10 and *w*_*A*_ = 3 *×* 10^−4^; (B) *n* = 3 and *w*_*A*_ = 3 *×* 10^−4^; (C) *n* = 10 and *w*_*A*_ = 5 *×* 10^−5^; and (D) *n* = 3 and *w*_*A*_ = 5*×*10^−5^. Simulations are initialised based on the UK data on 1st September 2020 with 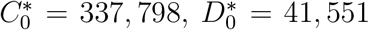 and model parameters are *α*_0_ = 0.05 [days^−^], *α* = 0.4 [days], *β* = 0.07 [days^−1^], *γ* = 0.05 [days^−1^], *δ* = 0.03 [days^−1^], *ρ* = 0.05, *η* = 0.1 [days^−1^], *κ* = 10, *ζ* = 0.15, and *T*_*d*_ = 92 [days] corresponding to the 2nd of December 2020. The *τ* –leaping approximate stochastic simulation algorithm is used with time-step *τ* = 1 [days].

#### 2.2.2 Incorporation of two-dose vaccination program

We now extend the base model described in Section 2.2.1 to incorporate vaccinations. We consider only a single vaccine-brand (e.g., AstraZeneca or Pfizer only) with a two dose vaccination protocol. These restrictions substantially reduce the complexity of our model and we highlight in Section 4 some implications for use with more complex situations with multiple vaccines and boosters. To implement the dynamics of vaccinations, we substitute each compartment in the base model with three new compartments representing vaccination status: unvaccinated, vaccinated (1st dose), and fully vaccinated (2nd dose) (Figure 4). For example, the susceptible compartment, *S*, in the base model becomes unvaccinated susceptible (*S*_*u*_), vaccinated susceptible (*S*_1_) and fully vaccinated susceptible (*S*_2_) (See Table 1).

**Figure 4.**
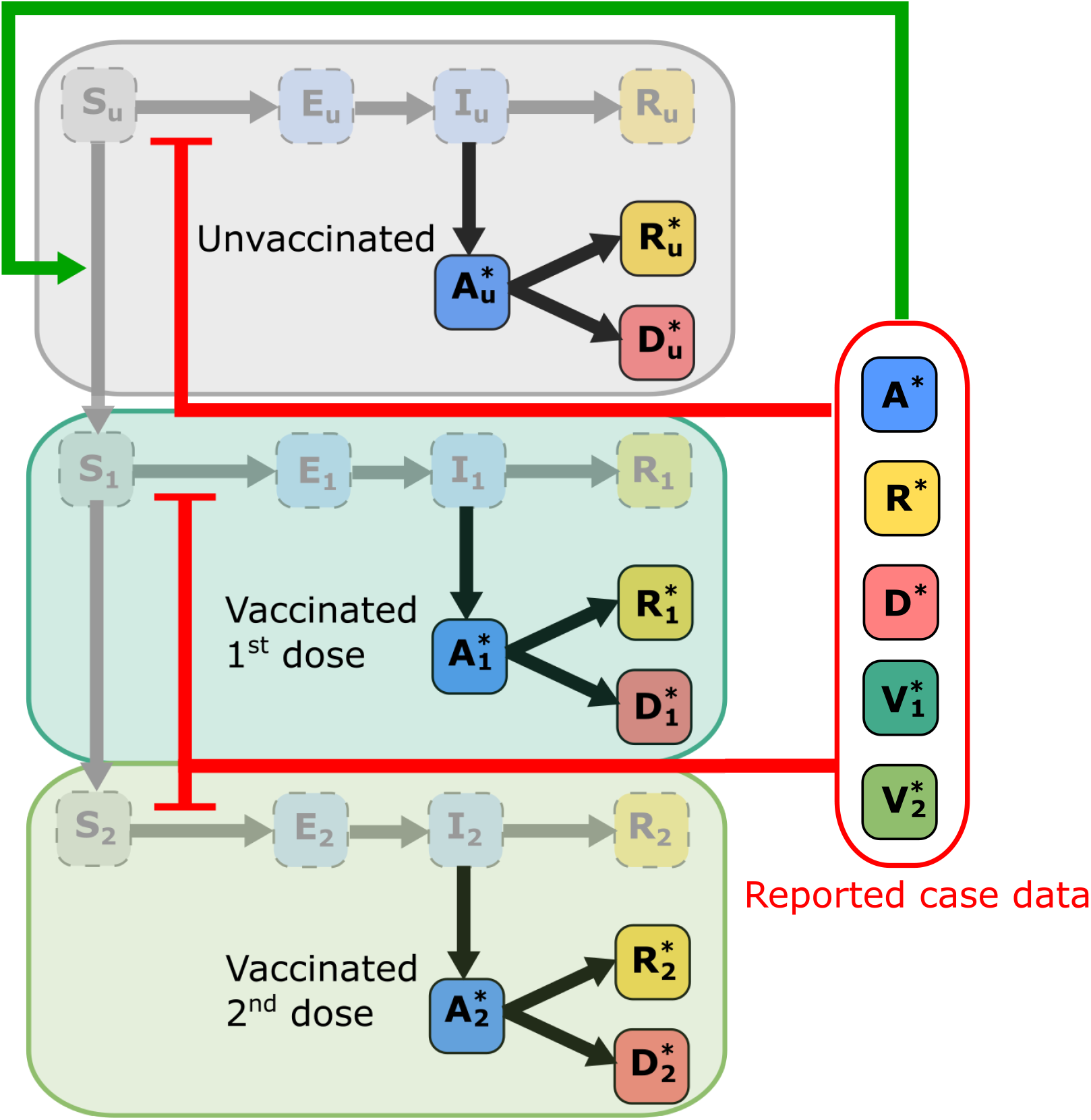
Schematic of the epidemiological model including two-dose vaccination program. States are marked as labelled boxed and arrows indicate state transitions. The model consists of three vaccinations stages, each with its own SEIR model with the reported case data arising through the transition from *I*_*u*_ to 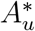 (resp. *I*_1_ to 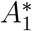 pr *I*_2_ to 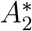). The transmission is inhibited by a response function that depends on the aggregated observed reported case data. Similarly, vaccination is promoted thought the hesitancy effect function that depends on the aggregated observed reported case data.

We assume that those currently infected with COVID-19, that is exposed and infectious individuals, do not get vaccinated until after recovery. This simplifies the model so that the only possible transitions between vaccination levels are for the susceptible and recovered compartments, *S*_*u*_, *R*_*u*_, and 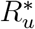. Importantly, here the observable states are different since available data sources only report total counts for cases, deaths and vaccinations. Therefore we denote five observables, the total confirmed active cases 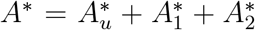, total case deaths, 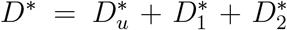, total case recoveries, 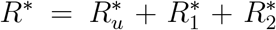, vaccinations (1st dose), 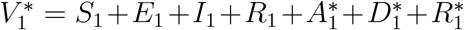, and fully vaccinated 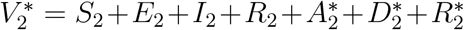 (Figure 4).

At each level *i* ∈ {1, 2} of vaccination, modifiers 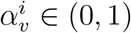 and 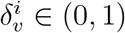 are applied to the transmission and death rates, respectively. That is, at vaccination level *i* the effective transmission rate is 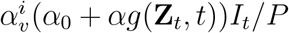 and the effective death rate is 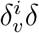 (Figure 4). Estimates for these parameters can be obtained from the literature on the efficacies of the particular vaccine of interest at each level of vaccination (Lopez Bernal et al., 2021a,b; Rosenberg et al., 2021; Voysey et al., 2020). For example, for the Oxford-AstraZeneca vaccine (ChAdOx1) against the B.1.617.2 (Delta) variant we can obtain values 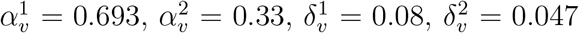, and 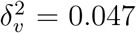 (Lopez Bernal et al., 2021b). There are two vaccination rate parameters *ν* > 0 and *ω* > 0, with *ν* representing the maximum possible vaccination rate in the absence of any vaccine hesitancy, and *ω* is the rate from first dose to second dose as per protocol. Figure 5 demonstrates the effect that vaccination has on the virus spread and severity under different scenarios of response behaviour and under the assumption of perfect vaccination rate of *ν*. For the continued lockdown protocol case Figure 5(A),(C) and the case of ending lockdowns once vaccines are available Figure 5(B),(D). We observe, as expected, a reduction it the total case deaths due to vaccination (Compare Figure 5(A) with Figure 5(C) and Figure 5(B) with Figure 5(D)). We also note a change of elimination occurring when restrictions continue along with the vaccination program (Figure 5(C)).

**Figure 5.**
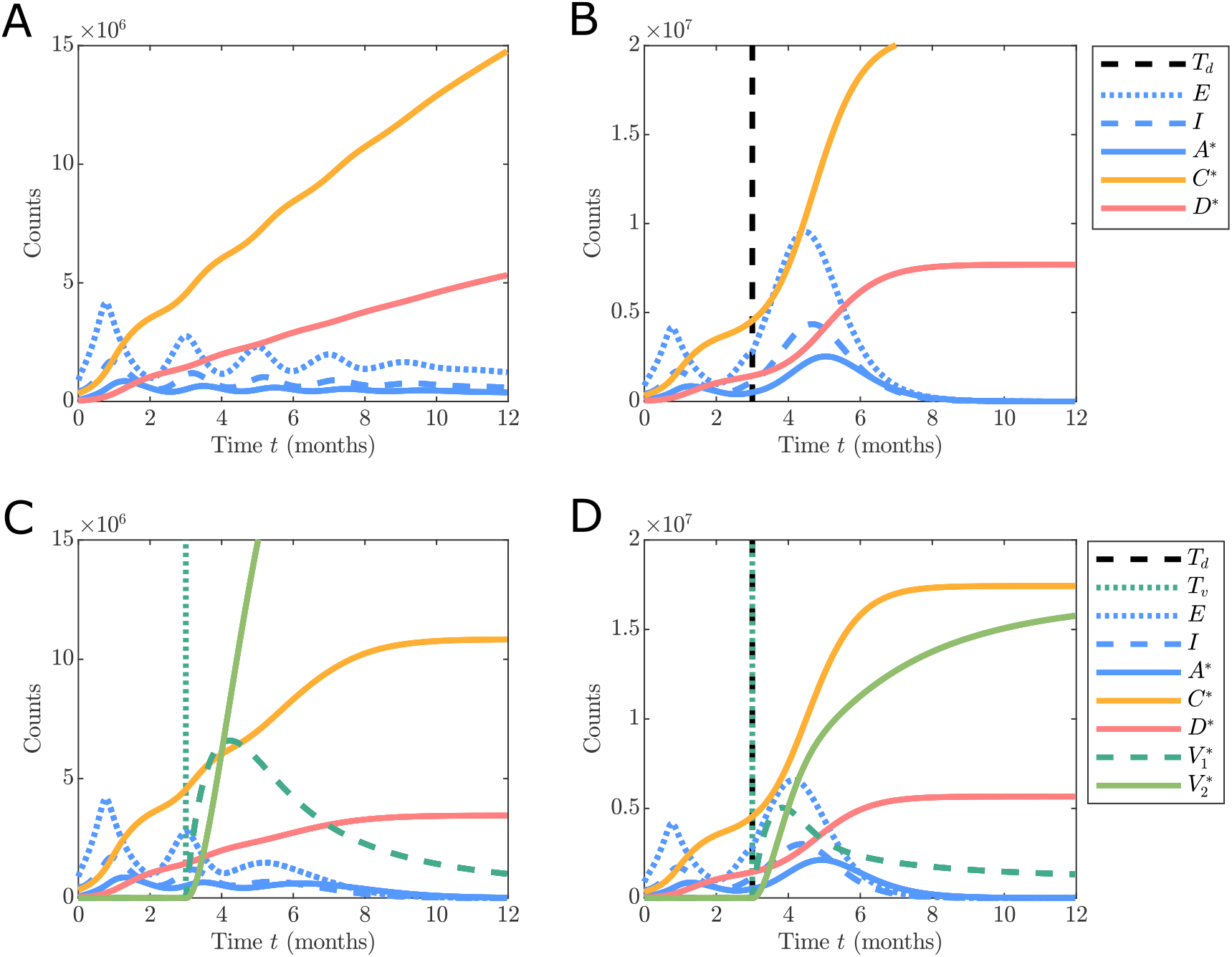
Comparison of the base epidemiological model (A)–(B) with the vaccination model (C)–(D) under different configurations response function and vaccination inclusion (in the absence of hesitancy effects). (A) and (C): *g*(*A*\*, *t*) = 1*/*(1 + (*w*_*A*_*A*\*)^*n*^), (B) and (D): 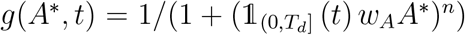. Simulations are initialised based on the UK data on 1^st^ September 2020 with 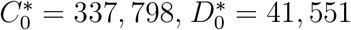 and model parameters are *α*_0_ = 0.05 [days^−1^] *α* = 0.4 [days^−1^], *β* = 0.07 [days^−1^], *γ* = 0.05 [days^−1^], *δ* = 0.03 [days^−1^], *ρ* = 0.05, *η* = 0.1 [days^−1^], *κ* = 10 [−], *ζ* = 0.1527 [ − ], *n* = 10, *w*_*A*_ = 5 *×* 10^−5^, *T*_*d*_ = 92 [days], *ν* = 0.01, *ω* = 0.048, 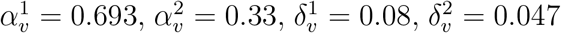, and *T*_*v*_ = 92 [days] corresponding to the 2nd of December 2020. The *τ*-leaping approximate stochastic simulation algorithm is used with time-step *τ* = 1 [days].

Vaccine hesitancy is incorporated through another feedback loop: we denote as the *hesitancy effect* function *h*(·) ∈ [0, 1] resulting in an effective vaccination rate of *νh*(·) (Figure 4). We assume that vaccine hesitancy behaviour only affects the probability of an individual receiving the first dose with every individual that gets vaccinated continuing to obtain a second dose to be “fully vaccinate”. We note that this assumption would not be appropriate for subsequent boosters, as there is the major concern of vaccine fatigue in relation to boosters. Therefore extensions to the model to account for boosters should model hesitancy behaviours for boosters also. Just as with the response function, the hesitancy effect function only depends on the observable states and time. We treat *h*(·) as an increasing function with *h*(·) = 0 corresponding to a situation where individuals refuse vaccination entirely and *h*(·) = 1 leads to no hesitancy effect with the maximum rate *ν* achieved. This function is intended to model the way in which reported case and vaccination data may increase the probability of a individual seeking vaccination. It is well established that the incidence of a disease in a community will tend to increase the likelihood of an individual to seek vaccination (Buonomo, 2020; Buonomo et al., 2022). If there are no vaccination safety concerns in the community, any delays in vaccine uptake may be due to complacency in regions where there was initially few COVID-19 outbreaks (such as Australia and New Zealand). This could be modelled with the function,

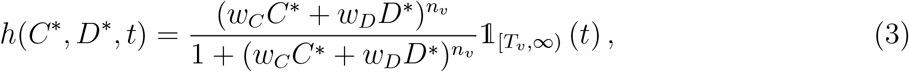

where *w*_*C*_, *w*_*D*_ > 0 are weights and *n*_*v*_ > 0 slope parameter, and *T*_*v*_ is that time the vaccine roll-out starts with 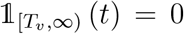 for *t < T*_*v*_. The relative importance of case number and deaths in influencing an individuals decision to get vaccinated cane be assessed by considering the case where *C*\* = (*w*_*D*_*/w*_*C*_)*D*\*, that is each new death has as much influence as *w*_*D*_*/w*_*C*_ new cases. Of course, motivation could be completely dominated by cases (resp. deaths), in which cases *w*_*D*_ = 0 (resp. *w*_*C*_ = 0). Another cause of vaccine hesitancy behaviour are vaccine safety concerns which were more prevalent in the COVID-19 pandemic due to rapid development of vaccines (Dror et al., 2020) and some widely publicised, though rare, side effects (Lau et al., 2021; Sinclair et al., 2022). If only vaccination safety is a concern, then critical mass of fully vaccinated individuals may be necessary to alleviate these concerns. This can be reflected in the function,

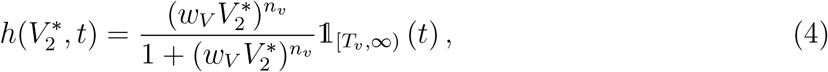

with weight parameter *w*_*V*_ > 0. Note that in this case, at time *T*_*v*_ some initial non-zero vaccinated population must be present, otherwise the vaccinated population will remain at 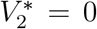. The initial population could represent participants in relevant clinical trials, or an initial vaccination mandate for particular occupations (e.g. healthcare workers). Finally, we can consider a population hesitancy behaviour that is due to safety concerns and complacency,

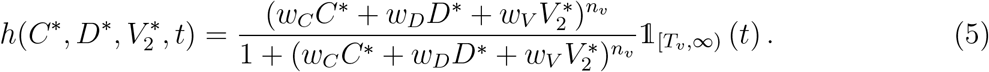

Once again the weights *w*_*C*_, *w*_*D*_, *w*_*V*_ ≥ 0 provide a measure of the relative influence each new case, death or vaccination has on he probability of an individual getting vaccinated. In reality, these parameters will be unknown, however if standard COVID-19 case data can provide insight into the values of this parameters, then we can start to assess the trends in hesitancy behaviours for a given population of interest.

### 2.3 Simulation study

Given that reported data is aggregated, it is unclear if the data are informative enough to identify what type of hesitancy effect is occurring in reality. For example, three different scenarios are shown in Figure 6 corresponding to no hesitancy (Figure 6(A)), complacency only (Figure 6(B)), and complacency and vaccine safety concerns (Figure 6(C)). While each evolution is different qualitatively, it is completely unclear if the scenarios including different hesitancy effects can be distinguished from a scenario with no hesitancy effect but slower vaccination rate. For example, such is the difference in the hesitancy behaviour between Figure 6(A) and Figure 6(B), yet the differences in the dynamics are subtle.

**Figure 6.**
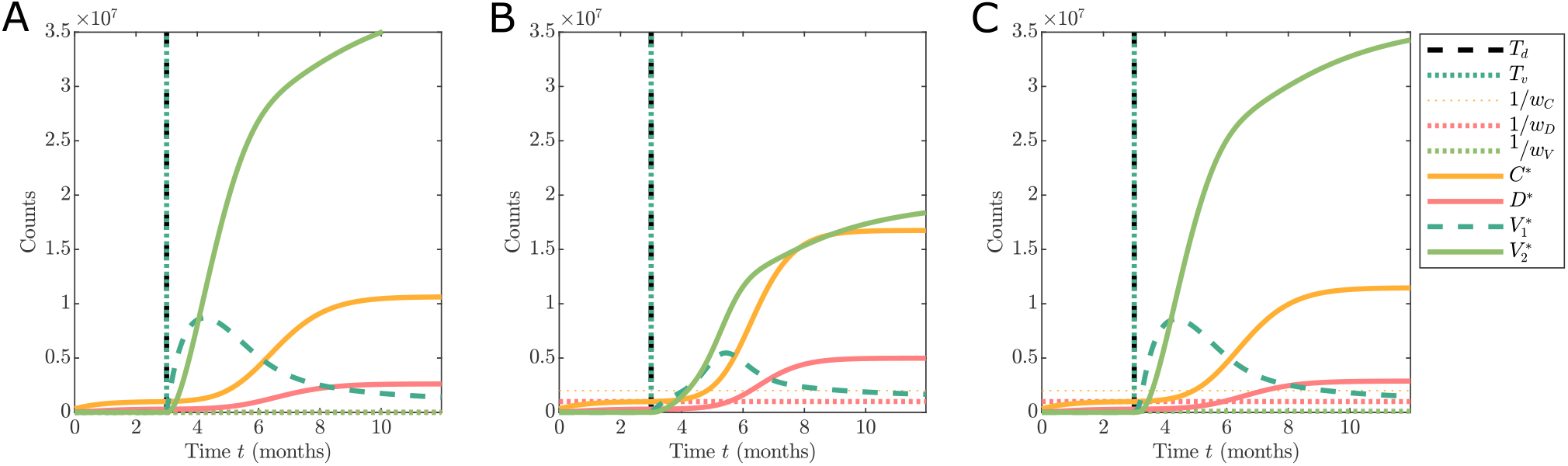
Comparison of different hesitancy effect functions show the dynamics of the observable states. The scenarios are: (A) no hesitancy behaviour with 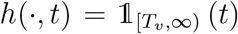; (B) hesitancy effect driven by complacency only with *h*(·) defined in Equation 3; and (C) hesitancy effect driven by mild safety concerns and some complacency only with *h*(·) defined in Equation 5. Simulations are initialised based on the UK data on 1st September 2020 with 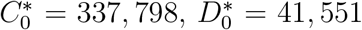, and model parameters are 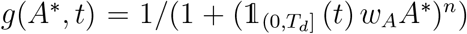, *α*_0_ = 0.05 [days^−1^], *α* = 0.4 [days^−1^], *β* = 0.07 [days^−1^], *γ* = 0.05 [days^−1^], *δ* = 0.03 [days^−1^], *ρ* = 0.05, *η* = 0.1 [days^−1^], *κ* = 10, *ζ* = 0.15, *n* = 10, *w*_*A*_ = 3 *×* 10^−4^, *T*_*d*_ = 92 [days], *ν* = 0.01, *ω* = 0.048, 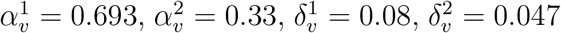, *n*_*v*_ = 4, *w*_*C*_ = 2 *×* 10^−6^, *w*_*C*_ = 1 *×* 10^−6^, *w*_*C*_ = 1 *×* 10^−5^, and *T*_*v*_ = 92 [days] corresponding to the 2nd of December 2020. The *τ*-leaping approximate stochastic simulation algorithm is used with time-step *τ* = 1 [days].

The behaviour observed in Figure 6 provides motivation to explore the informativity of aggregated reported COVID-19 data as provided by online repositories for the purpose of analysing vaccine hesitancy behaviour. To explore this, we consider a simulation study and develop Bayesian analysis techniques.

We generate multiple simulated datasets under three different vaccine uptake behaviours. Specifically, we consider: complacency-based hesitancy (*w*_*V*_ = 0, *w*_*C*_ > 0, and *w*_*D*_ > 0), complacency and vaccine safety (*w*_*V*_ > 0, *w*_*C*_ > 0, and *w*_*D*_ > 0) hesitancy, and no hesitancy (*h*(·) = 1). In each of the simulation settings we vary the weights and the maximum vaccination rate, the remaining parameters are fixed based on the literature (Lopez Bernal et al., 2021a,b; Rosenberg et al., 2021; Voysey et al., 2020; Warne et al., 2020) as listed in Table 2.

### 2.4 Bayesian analysis

Give a simulated dataset, 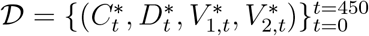, we infer the parameters related to the vaccination uptake ***θ*** = [*ν, n*_*v*_, *w*_*C*_, *w*_*D*_, *w*_*V*_] through sampling the Bayesian posterior distribution with density,

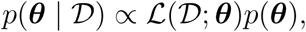

where ℒ (***θ***; 𝒟) is the likelihood function and *p*(***θ***) is the prior probability density. Due to the stochastic nature of the model and the large number of latent states, the likelihood function is intractable, and we rely upon approximate Bayesian computation (ABC) (Sisson et al., 2018; Sunnåker et al., 2013) to sample the approximate posterior,

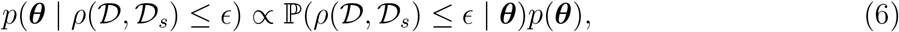

where 𝒟_*s*_ is a simulated dataset generated from the model with given parameter values, ***θ***, *ρ*(𝒟, *𝒟*_*s*_) is a discrepancy metric between the data and simulations, and *ϵ* is a target discrepancy threshold. For this work, our discrepancy metric between two datasets is the Frobenius norm of the matrix of differences between counts in each dataset (Supplementary Material). To achieve accurate inferences we apply an adaptive sequential Monte Carlo sampler for ABC (SMC-ABC) (Drovandi and Pettitt, 2011) that produces *N* samples with as small a discrepancy as practically possible. We repeat this inference process for all *M* synthetic datasets, 𝒟_1_, 𝒟_2_, …, 𝒟_*M*_, that span the different hesitancy scenarios. Thereby arriving at *M* sets of posterior samples 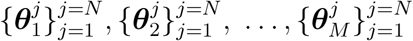. The structure of the resulting posterior samples can be used to investigate if there is information about vaccine hesitancy available in COVID-19 case data and vaccination counts. In all cases the priors are *ν* ∼ *𝒰* (0, 0.02), *n*_*v*_ ∼𝒰 (0, 10), − log *w*_*C*_ ∼𝒰 (3, 8), −log *w*_*D*_ ∼𝒰 (3, 8), and −log *w*_*V*_ ∼𝒰 (3, 8).

Model fit is assessed through sampling the within-sample posterior predictive distribution with density,

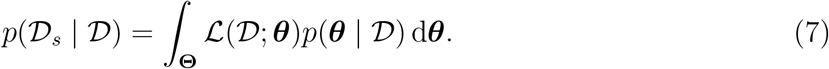

If the model fits well, then we expect the data to lie within the 95% Credible Intervals (95% CrI). Since the models we use to generate synthetic data are all special cases of the full model that is based on Equation (5) and Equation (6), there is no model misspecification error to account for in the inference. However, the posterior predictive check is useful to assess the accuracy of the SMC-ABC sampler with respect to the discrepancy threshold.

### 2.5 Identifiability analysis

In the setting of deterministic models, a parameter is defined as *structurally identifiable* if the mapping from parameter value, to a model solution curve is an invertible function. Various methods to analyse structural identifiability for deterministic dynamical systems are available in the literature (Bellu et al., 2007; Chiş et al., 2011a,b; Ligon et al., 2017). In contrast, very few methods are available for structural identifiability for stochastic models (Browning et al., 2020; Pieschner et al., 2022). For simplicity, we derive a deterministic ordinary differential equation (ODE) version of our model presented in Section 2.2.2 and apply generating series approaches to investigate identifiability (Ligon et al., 2017). This is appropriate in this setting as the ODE can be considered as an approximation of the mean of the stochastic process as the variance is comparatively negligible due to the large populations in each compartment (Browning et al., 2020).

In addition to structural identifiability, Bayesian posterior samples provide a way to explore practical identifiability. While frequentist approaches exist for exploring practical identifiability, such as profile likelihoods (Pawitan, 2001; Murphy et al., 2022; Simpson et al., 2022), they are challenging to apply for models with intractable likelihood functions. Therefore we adopt a Bayesian approach where practical identifiability is primarily explored via the marginal posterior distributions typically using Markov chain Monte Carlo (Browning et al., 2020; Hines et al., 2014) and posterior predictive sampling. Since we are sampling from the joint posterior distribution we automatically have access to samples from the marginal posterior distributions. When marginals themselves demonstrate practical non-identifiability, exploring correlation structures can provide additional insight. In this work, we visually inspect the bivariate marginal structures for each simulation case and Spearman’s rank correlation between pairs of parameters.

## 3. Results

In this section, we present the results of our simulation study. We highlight the key simulation scenarios explored and appropriateness of the model calibration process. We then demonstrate that while individual vaccine uptake parameters may not be uniquely identified from COVID-19 case data, the posterior distributions contain sufficient information to determine the vaccination scenario that is occurring in the data.

### 3.1 Simulation scenarios and model calibration

Synthetic datasets were constructed using the full model (Section 2.2.2) under each vaccination hesitancy behaviour scenario considered in Section 2.3. Example synthetic datasets for these three cases are shown in Figure 7.

**Figure 7.**
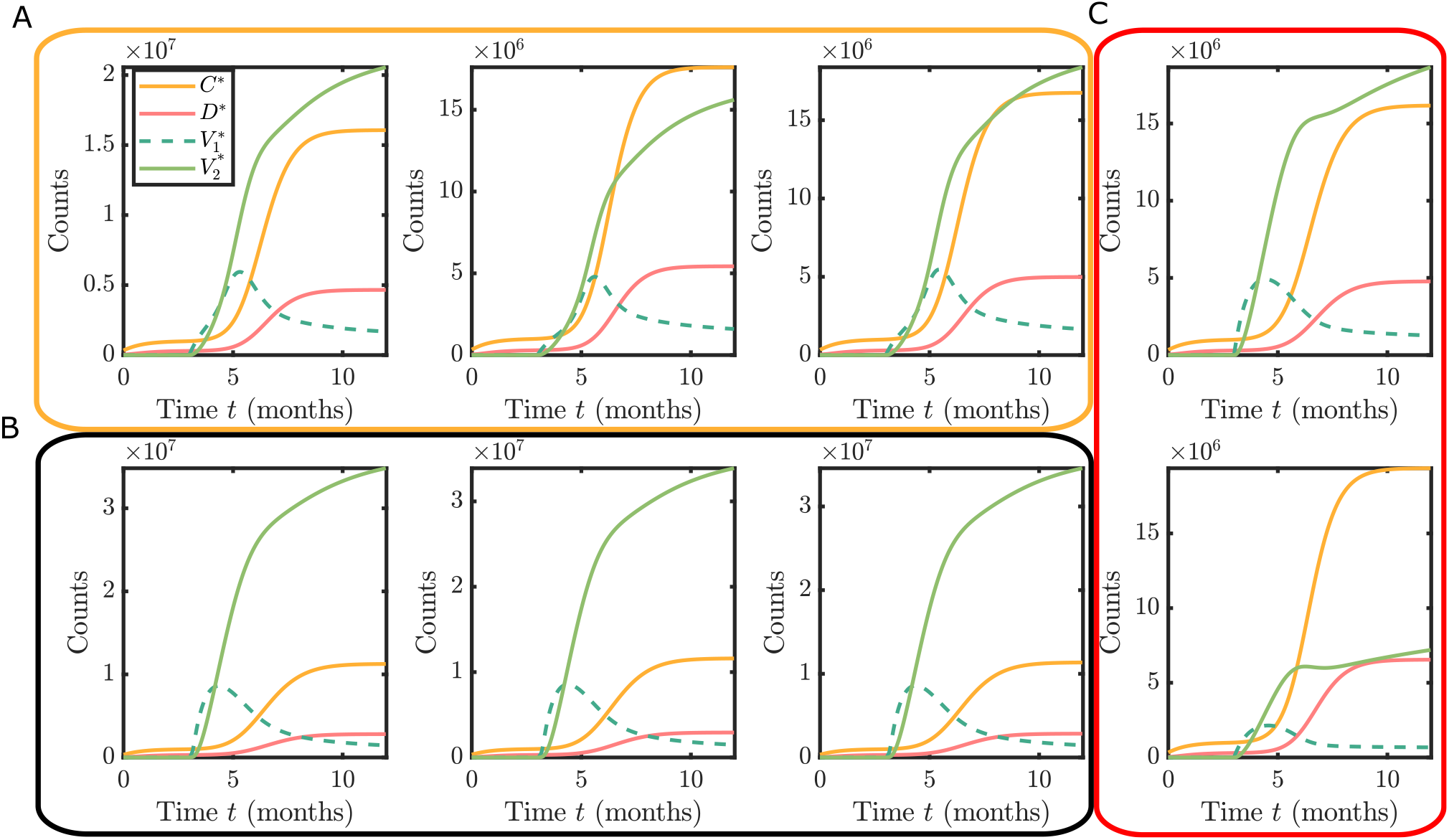
Example synthetic datasets for the three hesitancy scenarios: (A) complacency only with *w*_*C*_ > 0, *w*_*D*_ > 0 and *w*_*V*_ = 0; (B) complacency and vaccine safety concerns with *w*_*C*_ > 0, *w*_*D*_ > 0 and *w*_*V*_ > 0; and (C) no hesitancy, but smaller maximum vaccination rate than in (A) and (B).

For each dataset, 𝒟, we apply adaptive SMC-ABC (Supplementary Material) targeting the joint posterior of the vaccination uptake parameters *p*(*ν, n*_*v*_, *w*_*C*_, *w*_*D*_, *w*_*V*_ | 𝒟). The result is *N* = 1000 samples from the approximate posterior for each synthetic dataset. These samples are used for Monte Carlo integration of posterior probability densities, practical identifiability analysis, and visualisation of parameter correlation structures to investigate the use of COVID-19 case data for identification of vaccine hesitancy patterns.

The model fitness is evaluated for each synthetic dataset using within-sample posterior predictive checks. Since the model used for each synthetic dataset is nested within our full model we do not need to account for model misspecification. The purpose of the posterior predictive check is to ensure the sampler has adapted to a sufficiently small discrepancy threshold for inference. Figure 8 provide examples posterior predictive simulations and demonstrates typical model fit obtained from the Bayesian calibration using equivalent convergence criteria for the posterior sampler. Interestingly, the synthetic data scenarios based on complacency hesitancy behaviour only demonstrate a greater computational challenge for calibration with larger uncertainty intervals in the posterior predictive distribution (Figure 8B). Conversely, the synthetic data scenarios based on complacency and vaccine safety hesitancy (Figure 8A) or no hesitancy show (Figure 8C) much narrower credible intervals.

**Figure 8:**
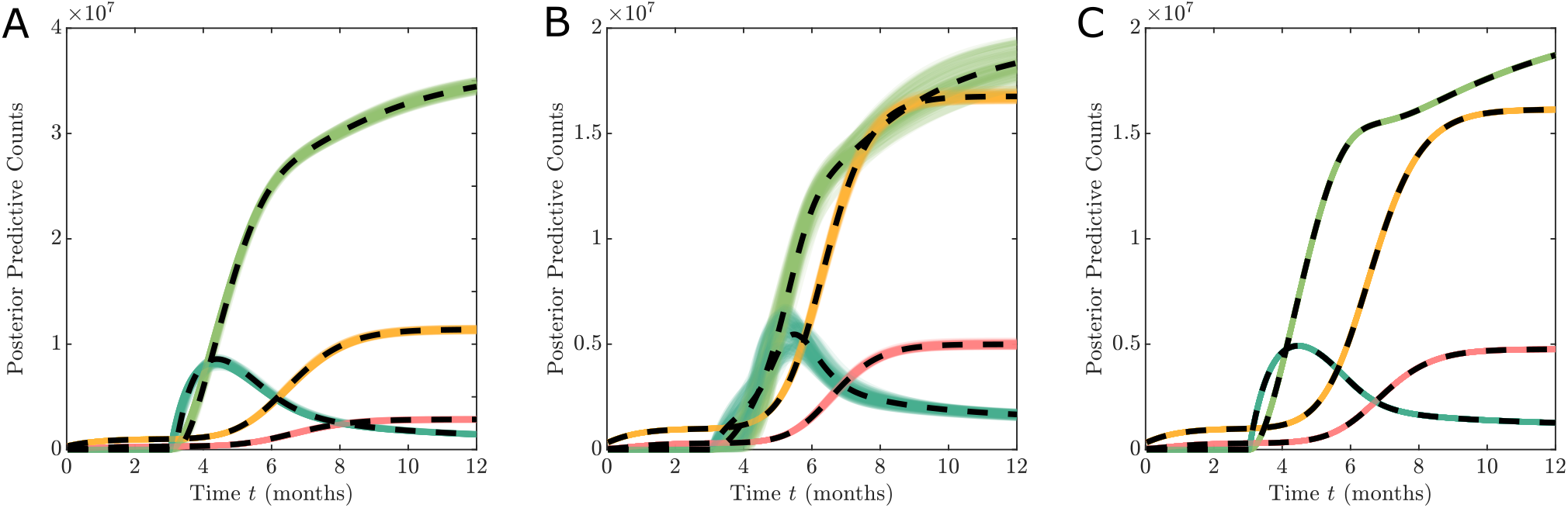
Example posterior predictive simulations (shaded regions) demonstrating model fit for different synthetic datasets (black dashed lines). (A) Complacency and vaccine safety concerns (true parameters ***θ*** = (0.01, 4, 5 *×* 10^−7^, 1 *×* 10^−6^, 1 × 10^−5^)), (B) complacency only (true parameters ***θ*** = (0.01, 4, 5 *×* 10^−7^, 1 *×* 10^−6^, 0)), and (C) no hesitancy (true parameters ***θ*** = (0.005, 4, 1, 1, 1)).

### 3.2 Identifiability of hesitancy function parameters

We use the GenSSI 2.0 toolkit (Chiş et al., 2011a; Ligon et al., 2017) to analyse the structural identifiability of the parameters related to vaccination uptake ***θ*** = (*ν, n*_*v*_, *w*_*C*_, *w*_*R*_, *w*_*V*_). This approach is applied to a deterministic version of the model developed in Section 2.2.2 for the multi-phase response function (Equation (2)) and hesitancy effect function (Equation (5)). We consider the case when *T*_*v*_ = *T*_*d*_, that is, NPI based restrictions are lifted at the same time as the vaccine roll-out, and the case when *T*_*v*_ ≪ *T*_*d*_, that is, NPI restrictions continue throughout the vaccination roll-out. In both cases we observe only the total reported case numbers, deaths, and vaccinations.

In both cases, all of the vaccine uptake parameters are shown to be locally structurally identifiable. This means that model parameters can be uniquely determined within as subset of parameter space. While this structural identifiability analysis is not directly applicable to the stochastic model, for very large case numbers, the stochastic effects are negligible. Therefore, we conclude that the observation process, if it was continuous, is informative enough to investigate hesitancy behaviours.

In addition, since our real observation process only records daily counts, we still need to explore practical identifiability as the data are not continuous noise-free observations. The posterior marginal distributions demonstrate practical non-identifiability. This can be observed in the example marginal posterior density plots (Figure 9). Regardless of the synthetic data scenario, the hesitancy function parameters *w*_*C*_, *w*_*R*_, and *w*_*V*_ show substantial uncertainty. The only parameter that is consistently identified is the maximum vaccination rate *ν*. While the shape of the marginals (particularly the skewness) provide some hints toward recovering the true hesitancy scenario for the respective synthetic data, the differences between the scenarios are subtle.

**Figure 9.**
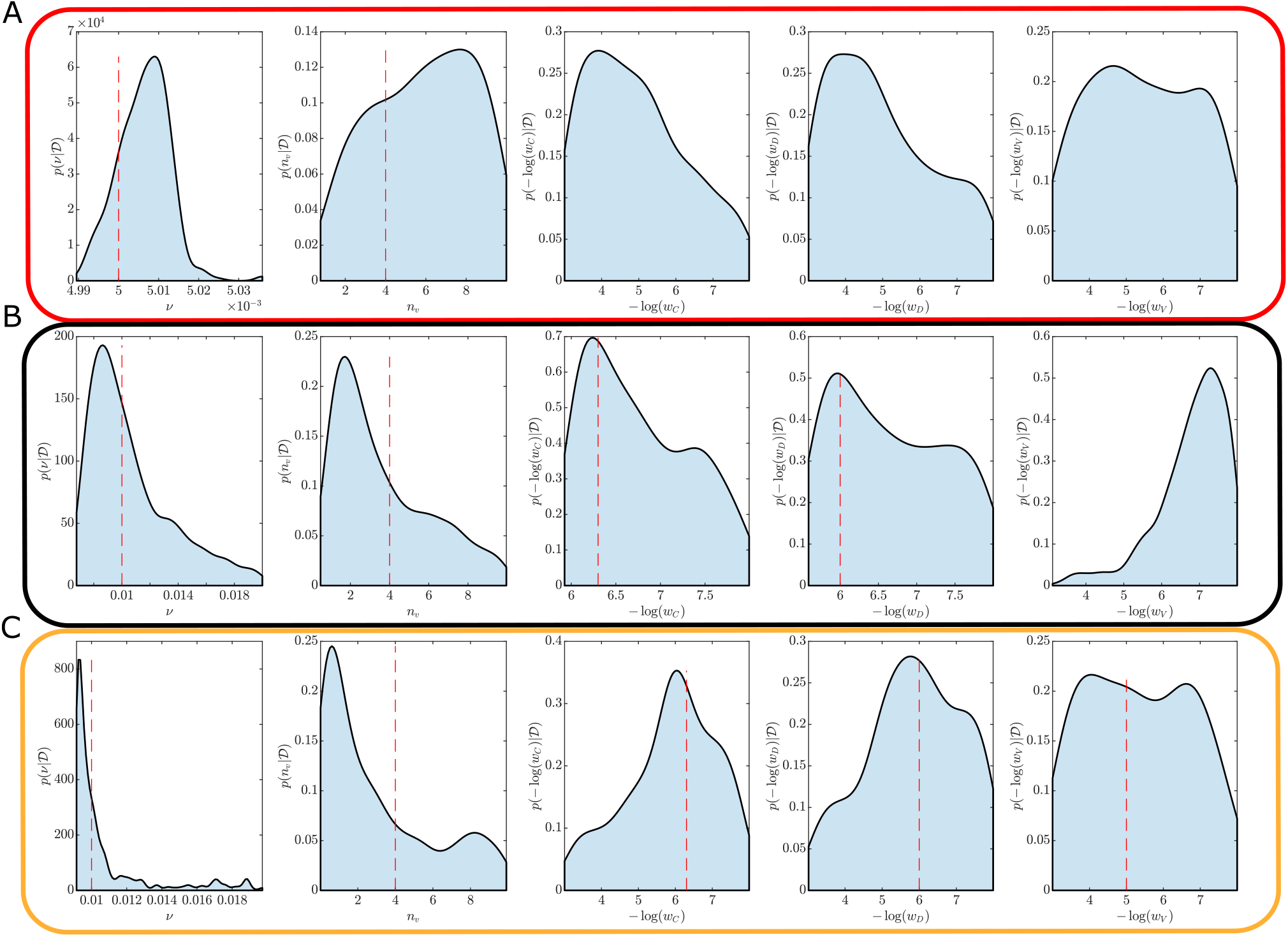
Example marginal posterior densities obtained using synthetic datasets for the three hesitancy scenarios: (A) no hesitancy; (B) complacency only; and (C) complacency and vaccine safety concerns. True parameter values are indicated (red dashed lines). Substantial uncertainties in the hesitancy function parameters *n*_*v*_, *w*_*C*_, *w*_*D*_,and *w*_*V*_ indicate practical unidentifiability problems. Cases without a dashed red line indicate cases when the true parameter corresponds to: (A) *w*_*C*_, *w*_*D*_, *w*_*V*_ → ∞; and (B) *w*_*V*_ → 0.

### 3.3 Hesitancy inferred through dependency structures

The marginal posterior distributions indicate that key parameters related to vaccination rates and hesitancy behaviours are practically non-identifiable using COVID-19 case data. However, practical non-identifiability does not preclude the study of the posterior structure to provide insight into the hesitancy behaviours. In particular, the joint posterior distributions contain information about the correlation structure between these parameters. The qualitative form of these correlation structures can be used to uncover the true hesitancy behaviour, albeit without the ability to quantify the actual parameter values reliably. Figures 10–12 show characteristic example bivariate plot matrices for each of the different vaccination hesitancy scenarios described in Section 2.3. Throughout, all statistical hypothesis tests are performed at the 0.01 significance level, and correlations refer to Spearman’s rank correlation.

**Figure 10.**
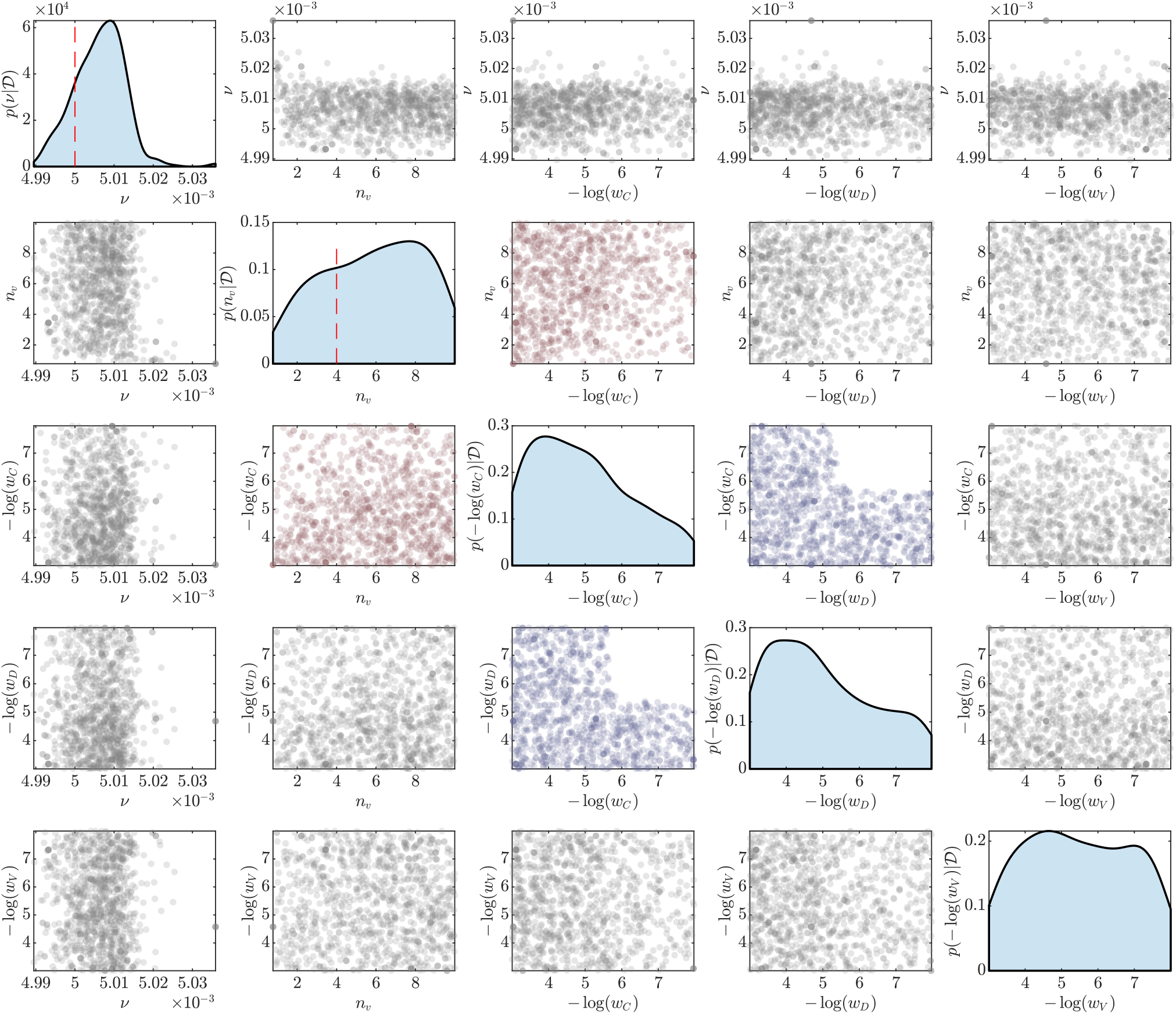
Example of posterior samples for the simulation scenario without any vaccine hesitancy. The main diagonals show marginal posterior densities with dashed red lines indicating the true values. Marginals without a dashed red line indicates the true parameter corresponds to *w*_*C*_, *w*_*D*_, *w*_*V*_ → ∞. The off-diagonal plots show the bivariate posterior samples, from which the pairwise correlations are computed. Plots with significantly positive, significantly negative and statistically insignificant correlations are shown in red, blue and grey respectively.

Firstly, by comparing Figure 10, that corresponds to synthetic data generated with no hesitancy, with Figures 11 and 12, that correspond to different hesitancy scenarios, we observe qualitative and quantitative differences in the bivariate posteriors. In the absence of hesitancy in the data, there is no evidence for correlation between *ν* and the negative log weights, or with the slope parameter *n*_*v*_ as all correlations are statistically insignificant (Figure 10). In contrast, there are statistically significant negative correlations between *ν* and *n*_*v*_, and statistically significant positive correlations between *ν* and negative log weights −log(*w*_*C*_), −log(*w*_*D*_), −log(*w*_*V*_) (Figures 11 and 12). This demonstrates that correlation between the maximum vaccinate rate *ν* with the negative log weight parameters −log(*w*_*C*_), −log(*w*_*D*_), and −log(*w*_*V*_) provide a strong indicator for the presence of hesitancy behaviours.

**Figure 11.**
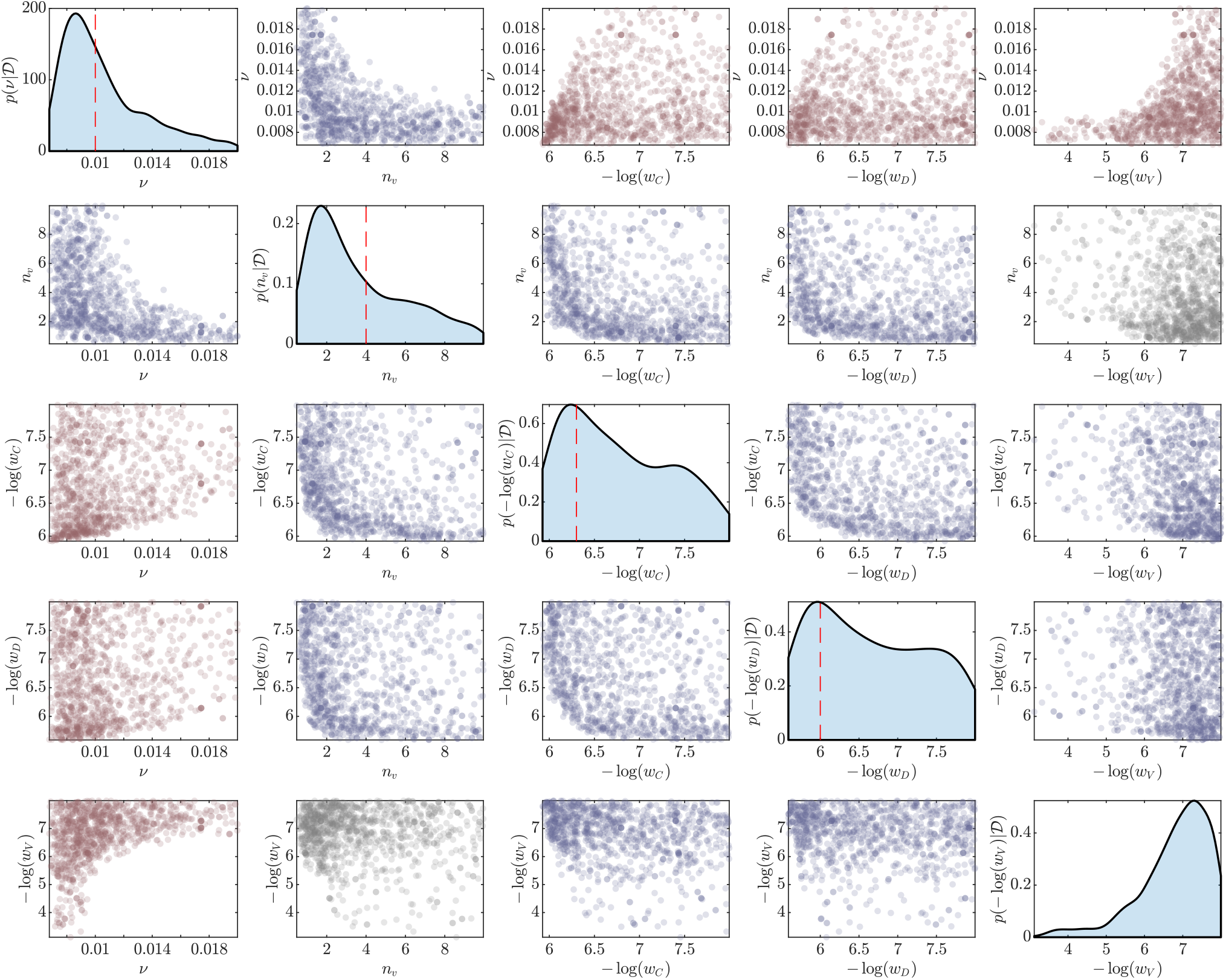
Example of posterior samples for the simulation scenario including complacency only. The main diagonals show marginal posterior densities with dashed red lines indicating the true values. The Marginal without a dashed red line indicates the true parameter corresponds to *w*_*V*_ → 0. The off-diagonal plots show the bivariate posterior samples, from which the pairwise correlations are computed. Plots with significantly positive, significantly negative and statistically insignificant correlations are shown in red, blue and grey respectively.

**Figure 12.**
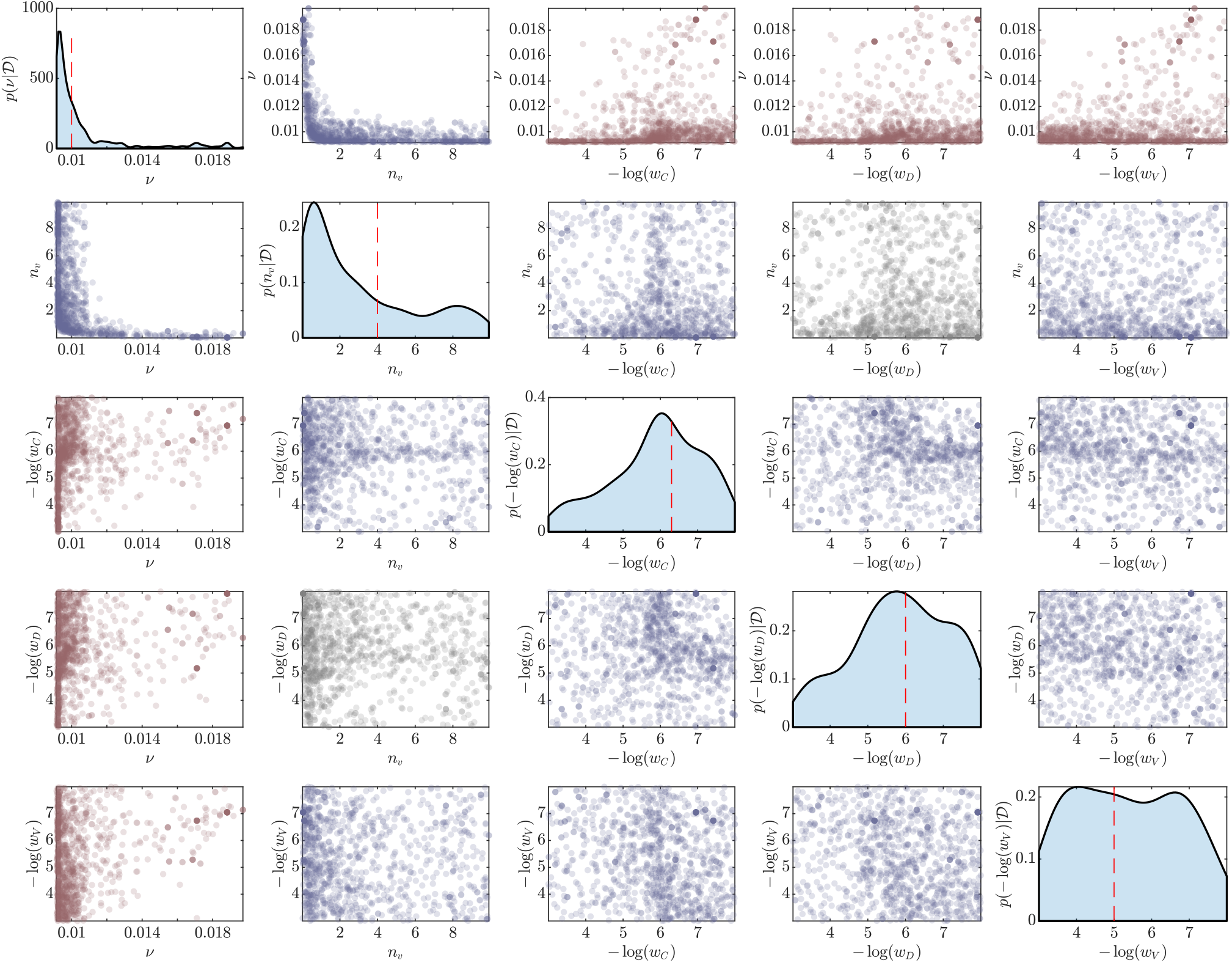
Example of posterior samples for the simulation scenario including complacency and vaccine safety concerns. The main diagonals show marginal posterior densities with dashed red lines indicating the true values. The off-diagonal plots show the bivariate posterior samples, from which the pairwise correlations are computed. Plots with significantly positive, significantly negative and statistically insignificant correlations are shown in red, blue and grey respectively.

Second, comparing correlation structures for different cases of hesitancy can inform the components that are dominating behaviour. For example, Figure 11 shows the results for a synthetic dataset with only complacency behaviour. Here, the strength of the correlation between *ν* and −log(*w*_*V*_) is substantially stronger than that of *ν* with −log(*w*_*C*_) and −log(*w*_*D*_). This indicates the vaccine counts have a much smaller effect on the vaccine uptake than the confirmed cases and deaths. By comparison, a scenario that includes both complacency and vaccine safety concerns (Figure 12) has weaker correlation strengths that are on similar scales. The interpretation here is that increased cases, deaths and vaccine counts all increase the probability of individuals get vaccinated.

Finally, the correlations between the slope *n*_*v*_ parameters also provide some insight, but this information mainly complements the insights obtained through correlations with *ν* and the other parameters. These correlations seem to allow some identification of the dominating complacency drivers between total case numbers or death numbers, however, more work is required to explore this in more detail.

### 3.4 Summary

Through simulated synthetic dataset scenarios and Bayesian analysis, we demonstrate the ability to identify the presence of vaccine hesitancy at a population level. This is possible despite practical non-identifiability of individual parameters. We achieve this though inspection of the correlation structures in the joint posterior distribution for the parameters relevant to vaccine hesitancy. Further, once vaccine hesitancy behaviours have been identified, it is possible to isolate the specific factors that are driving the hesitancy behaviour. We explore the potential implications and limitations of these results in the Discussion (Section 4)

## 4 Discussion

In this work, we explore to feasibility of isolating hesitancy behaviour and key drivers of this hesitancy using reported case data, such as those available from COVID-19 online dashboards. Using a stochastic epidemic model that accounts for the effects of NPIs and vaccinations over time, we generate synthetic datasets for various hesitancy scenarios with model parameterisation inspired by the early AstraZeneca COVID-19 vaccination roll-out in the United Kingdom. While hesitancy parameters are structurally identifiable, Bayesian analysis reveals that there are practical identifiability challenges for the stochastic model. Despite this, our analysis framework can distinguish the different hesitancy behaviours through inspection of the joint posterior densities and correlation structures. This is potentially a useful tool for future vaccination rollouts for many diseases aside from COVID-19.

This analysis implies that, in principle, protocols could be developed to quantify the population level effects of hesitancy behaviour during a vaccine roll-out. This could supplement traditional approaches based on survey data to develop information and education campaigns to target hesitancy causes that will have the greatest impact on vaccine uptake. This could be particularly valuable in supplementing survey data when the epidemic situation is rapidly changing as sudden changes in attitudes are difficult to capture with survey data (Lindvall and Rönnerstrand, 2022). Furthermore, if reported data is appropriately subdivided, differences in hesitancy behaviour could be assessed for different jurisdictions or vaccine eligibility groups. A particular example of where this might have been applicable was during the COVID-19 vaccine roll-out of AstraZeneca when various regulators adjusted the minimum age recommendations in response to concerns around thrombosis and thrombocytopenia syndrome (Lau et al., 2021; Leask et al., 2021).

There are several key assumptions within our framework and it is important to highlight them here and discuss the applicability of our methods for real data. Firstly, we consider a spatially homogeneous population in which all individuals have an equal probability of interaction. Therefore, the framework we present here is largely applicable to smaller spatial scales, such as cities rather that countries. Second, we do not consider an age-structured population nor a staged vaccination roll-out. This does not preclude the utility of our method to be applied to staged programs, provided the reported data is available for the various ages structures and eligibility criterion for each vaccine stage. Due to the dimensionality of the problem, we recommend investigating hesitancy within each eligibility sub-group independently. Third, we assume there is sufficient information on the epidemic and vaccine efficacies to obtain values for all parameters except the vaccination rate and hesitancy parameters. This is a relatively minor constraint, since effective vaccines are unlikely to be rolled out for a virus that has unknown transmissibility, recovery and fatality rates. Finally, we only consider a single vaccine with a two dose regime with no hesitancy effects on the second dose. However, the overall approach would still be applicable to multiple vaccine brands by either extending the model to incorporate more vaccination types, or treat vaccination efficacy parameters as an averaged effect. As many data sources do not provide detailed breakdowns of vaccination counts by brand, the latter approach may be preferred.

In this work we have focused on the identification of hesitancy behaviours for a single well-mixed population. As we have stated in our assumptions, this does not necessarily limit the use of our approach to more complex and realistic populations. However, future development of our methods could dramatically expand the rage of possibilities. Various extensions that account for spatial heterogeneity (Dutta et al., 2021), age-structure (Blyuss and Kyrychko, 2021), and seasonality (Merow and Urban, 2020) would improve the broad applicability of our methods to more realistic settings. As COVID-19 becomes endemic to a population effects of waning immunity and vaccination booster programs becomes important (Menni et al., 2022). Due to our focus on the initial roll-out of a new vaccine, we have not accounted for these effects here, however, future research should extend our approaches to this case. Finally, vaccine hesitancy could be different for each vaccine brand, therefore a multiple vaccine system could be developed, though it is unclear if the individual hesitancy behaviours will be identifiable in this case.

To implement any of the above mentioned improvements to our modelling framework requires either an extension of the compartment space or the inclusion of hierarchical effects. For models with intractable likelihood functions, as our stochastic model is, such extensions have the potential to dramatically increase the computational cost and inhibit the feasibility of including additional model complexity. However, advanced algorithms for likelihood-free Bayesian computation have the potential to resolve some of these issues. For example, the hierarchical modelling setting, Alahakoon et al. (2022) developed new computational schemes to deal with waning immunity in epidemiological modelling. Furthermore various approaches that exploit surrogate or approximate models have been shown to dramatically improve performance whilst sacrificing minimal accuracy (Bon et al., 2022; Everitt and Rowińska, 2020; Prangle, 2016; Prescott and Baker, 2020, 2021; Jasra et al., 2019; Warne et al., 2022a,b, 2018). Finally, state-of-the-art massively parallel computing hardware has potential to complement algorithmic advances to enable real-time analysis (Hurn et al., 2016; Kulkarni et al., 2020, 2022; Lee et al., 2010; Mahani and Sharabiani, 2015; Warne et al., 2022c).

## 5 Conclusions

Our modelling and computational framework provides a new approach to monitoring vaccine hesitancy using reported case data and vaccination counts that have been widely available during the COVID-19 pandemic. We show that vaccine hesitancy behaviours can be identified from these data provided sufficient information on the epidemic exists preceding the vaccine roll-out. A full Bayesian analysis is performed to be able to identify correlation structure in parameters. It is likely that vaccine hesitancy will continue to be a barrier for vaccine uptake and a major concern for governments during the roll-out of new vaccines in future epidemics. Therefore, our tools that enable the rapid assessment of trends in vaccine hesitancy behaviours within a population can greatly assist public health policy makers and practitioners in addressing public concerns.

## Data Availability

This is a simulation study, all data in for form of simulation outputs is available online at https://github.com/davidwarne/COVID-19-vax

## Acknowledgments

DJW and ALJ thank the Centre for Data Science at QUT for the First Byte Grant. CD was supported by an Australian Research Council Future Fellowship (FT210100260). AM received funding from the European Union’s Horizon 2020 research and innovation programme under grant agreement No 101016233. Computational resources were provided by the eResearch Office at QUT.

## Software Availability

Matlab implementations of models and analysis used in this work are available on GitHub https://github.com/davidwarne/vaccination-hesitancy-modelling.

## Footnotes

1 In the early period of the COVID-19 pandemic some dashboards did provide case recovery data, however, the counts were not as reliable as the reported cases and deaths.

