## Appendix for "Bayesian uncertainty quantification to identify population level vaccine hesitancy behaviours"

### Supplementary Material for "Bayesian uncertainty quantification to identify population level vaccine hesitancy behaviours"

David J. Warne*[1,2], Abhishek Varghese[1,2], Alexander P. Browning[3], Mario M. Krell[4], Christopher Drovandi[1,2], Wenbiao Hu[5], Antonietta Mira[6,7], Kerrie Mengersen[1,2], and Adrianne L. Jenner[1,2]

[1]School of Mathematical Sciences, Queensland University of Technology, Brisbane, Australia
[2]Centre for Data Science, Queensland University of Technology, Brisbane, Australia
[3]Mathematical Institute, University of Oxford, Oxford, UK
[4]Graphcore Inc., Palo Alto, USA
[5]School of Public Health and Social Work, Queensland University of Technology, Brisbane, Australia
[6]Institute of Computational Science, Universitá della Svizzera italiana, Lugano, Switzerland
[7]Dipartimento di Scienza e Alta Tecnologia, Universitá dell'Insubria, Varese, Italy

December 14, 2022

**Appendix C Additional Results** **7**

### Appendix A Stochastic model details

In this section, we present the mathematical details of the base stochastic epidemiological model and the extension including vaccinations and hesitancy behaviour.

#### A.1 Base Model Formulation

We implement our model as a stochastic compartmental epidemiological model consisting of 7 main compartment types (i.e., $S$ (susceptible), $E$ exposed, $I$ (infectious), $R$ recovered, $A^*$ (confirmed active case), $R^*$ (confirmed case recovery) and $D^*$ (confirmed case fatality)). We denote the state of the population at time $t$ as $\mathbf{X}_t = [S_t, E_t, I_t, R_t, A_t^*, D_t^*, R_t^*]^\mathrm{T}$. Under the assumption of a spatially homogeneous population and exponential waiting time between events, then we arrive at a discrete-state Markov process that can be expressed in the Kurtz random time-change representation,

$$\mathbf{X}_t = \mathbf{X}_0 + \sum_{j=1}^{M} Y_j \left( \int_0^t h_j(\mathbf{X}_s, s) \, \mathrm{d}s \right) \boldsymbol{\nu}_j, \tag{1}$$

where $M$ is the number of transition events, $\mathcal{E}_1, \mathcal{E}_2, \ldots, \mathcal{E}_M$, $h_j(\cdot)$ and $\boldsymbol{\nu}_j$ are, respectively, the hazard function and state change for event $\mathcal{E}_j$, and $Y_j(\cdot)$ is an inhomogeneous Poisson process with mean $\int_0^t h_j(\mathbf{X}_s, s) \, \mathrm{d}s$. For this model, we have $M = 6$ and

$$\begin{aligned}
&\mathcal{E}_1 : S \to E, && h_1(\mathbf{X}_t, t) = [\alpha_0 + \alpha g(\mathbf{Z}_t, t)] \frac{I_t S_t}{P}, && \boldsymbol{\nu}_1 = [-1, 1, 0, 0, 0, 0, 0]^\mathrm{T}, \\
&\mathcal{E}_2 : E \to I, && h_2(\mathbf{X}_t, t) = \beta E_t, && \boldsymbol{\nu}_2 = [0, -1, 1, 0, 0, 0, 0]^\mathrm{T}, \\
&\mathcal{E}_3 : I \to R, && h_3(\mathbf{X}_t, t) = \eta I_t, && \boldsymbol{\nu}_3 = [0, 0, -1, 1, 0, 0, 0]^\mathrm{T}, \\
&\mathcal{E}_4 : I \to A^*, && h_4(\mathbf{X}_t, t) = \gamma I_t, && \boldsymbol{\nu}_4 = [0, 0, -1, 0, 1, 0, 0]^\mathrm{T}, \\
&\mathcal{E}_5 : A^* \to D^*, && h_5(\mathbf{X}_t, t) = \delta A_t^*, && \boldsymbol{\nu}_5 = [0, 0, 0, 0, -1, 1, 0]^\mathrm{T}, \\
&\mathcal{E}_6 : A^* \to R^*, && h_6(\mathbf{X}_t, t) = \rho A_t^*, && \boldsymbol{\nu}_6 = [0, 0, 0, 0, -1, 0, 1]^\mathrm{T}.
\end{aligned}$$

Here, $\mathbf{Z}_t = [A_t^*, D_t^*, R_t^*]^\mathrm{T}$ refers to the vector of observables.

#### A.2 Full Model Formulation

We implement our model as a stochastic compartmental epidemiological model consisting of 21 the compartments, that consist of 7 main compartment types (i.e., $S$ (susceptible), $E$ exposed, $I$ (infectious), $R$ recovered, $A^*$ (confirmed active case), $R^*$ (confirmed case recovery) and $D^*$ (confirmed case fatality)) with each occurring in one of three levels of vaccinations status (e.g., for susceptible compartment: $S_u$ unvaccinated, $S_1$, vaccinated (1st dose), $S_2$ vaccinated (2nd dose)). We refer to the main manuscript for parameter and compartment definitions (Sections 2.1).

There are 28 transition events, $\mathcal{E}_1, \mathcal{E}_2, \ldots, \mathcal{E}_{28}$, between the 21 compartments. These transition events and their respective hazard functions, assuming a well mixed population

$P$, are

$$
\begin{aligned}
&\mathcal{E}_1 : \ S_u + I_u \to I_u + E_u, h_1(\mathbf{X}_t) = (\alpha_0 + \alpha g(\mathbf{Z}_t, t)) \frac{I_{u,t} S_{u,t}}{P}, \\
&\mathcal{E}_2 : \ S_u + I_1 \to I_1 + E_u, h_2(\mathbf{X}_t) = \alpha_v^1 (\alpha_0 + \alpha g(\mathbf{Z}_t, t)) \frac{I_{1,t} S_{u,t}}{P}, \\
&\mathcal{E}_3 : \ S_u + I_2 \to I_2 + E_u, h_3(\mathbf{X}_t) = \alpha_v^2 (\alpha_0 + \alpha g(\mathbf{Z}_t, t)) \frac{I_{2,t} S_{u,t}}{P}, \\
&\mathcal{E}_4 : \ E_u \to I_u, \qquad h_4(\mathbf{X}_t) = \beta E_u, \\
&\mathcal{E}_5 : \ I_u \to R_u, \qquad h_5(\mathbf{X}_t) = \eta I_u, \\
&\mathcal{E}_6 : \ I_u \to A_u^*, \qquad h_6(\mathbf{X}_t) = \gamma I_u, \\
&\mathcal{E}_7 : \ A_u^* \to R_u^*, \qquad h_7(\mathbf{X}_t) = \rho A_u^*, \\
&\mathcal{E}_8 : \ A_u^* \to D_u^*, \qquad h_8(\mathbf{X}_t) = \delta A_u^*, \\
&\mathcal{E}_9 : \ S_u \to S_1, \qquad h_9(\mathbf{X}_t) = \nu h(\mathbf{Z}_t, t) S_u, \\
&\mathcal{E}_{10} : R_u \to R_1, \qquad h_{10}(\mathbf{X}_t) = \nu h(\mathbf{Z}_t, t) R_u, \\
&\mathcal{E}_{11} : S_1 + I_u \to I_u + E_1, h_{11}(\mathbf{X}_t) = \alpha_v^1 (\alpha_0 + \alpha g(\mathbf{Z}_t, t)) \frac{I_{u,t} S_{1,t}}{P}, \\
&\mathcal{E}_{12} : S_1 + I_1 \to I_1 + E_1, \ h_{12}(\mathbf{X}_t) = (\alpha_v^1)^2 (\alpha_0 + \alpha g(\mathbf{Z}_t, t)) \frac{I_{1,t} S_{1,t}}{P}, \\
&\mathcal{E}_{13} : S_1 + I_2 \to I_2 + E_1, \ h_{13}(\mathbf{X}_t) = \alpha_v^1 \alpha_v^2 (\alpha_0 + \alpha g(\mathbf{Z}_t, t)) \frac{I_{2,t} S_{1,t}}{P}, \\
&\mathcal{E}_{14} : E_1 \to I_1, \qquad h_{14}(\mathbf{X}_t) = \beta E_1, \\
&\mathcal{E}_{15} : I_1 \to R_1, \qquad h_{15}(\mathbf{X}_t) = \eta I_1, \\
&\mathcal{E}_{16} : I_1 \to A_1^*, \qquad h_{16}(\mathbf{X}_t) = \gamma I_1, \\
&\mathcal{E}_{17} : A_1^* \to R_1^*, \qquad h_{17}(\mathbf{X}_t) = \rho A_1^*, \\
&\mathcal{E}_{18} : A_1^* \to D_1^*, \qquad h_{18}(\mathbf{X}_t) = \delta_v^1 \delta A_1^*,
\end{aligned}
$$

$$
\begin{aligned}
&\mathcal{E}_{19} : S_1 \to S_2, \qquad h_{19}(\mathbf{X}_t) = \omega S_1, \\
&\mathcal{E}_{20} : R_1 \to R_2, \qquad h_{20}(\mathbf{X}_t) = \omega R_1, \\
&\mathcal{E}_{21} : S_2 + I_u \to I_u + E_2, h_{21}(\mathbf{X}_t) = \alpha_v^2 (\alpha_0 + \alpha g(\mathbf{Z}_t, t)) \frac{I_{u,t} S_{2,t}}{P}, \\
&\mathcal{E}_{22} : S_2 + I_1 \to I_2 + E_1, h_{22}(\mathbf{X}_t) = \alpha_v^1 \alpha_v^2 (\alpha_0 + \alpha g(\mathbf{Z}_t, t)) \frac{I_{1,t} S_{2,t}}{P}, \\
&\mathcal{E}_{23} : S_2 + I_2 \to I_2 + E_2, h_{23}(\mathbf{X}_t) = (\alpha_v^2)^2 (\alpha_0 + \alpha g(\mathbf{Z}_t, t)) \frac{I_{2,t} S_{2,t}}{P}, \\
&\mathcal{E}_{24} : E_2 \to I_2, \qquad h_{24}(\mathbf{X}_t) = \beta E_2, \\
&\mathcal{E}_{25} : I_2 \to R_2, \qquad h_{25}(\mathbf{X}_t) = \eta I_2, \\
&\mathcal{E}_{26} : I_2 \to A_2^*, \qquad h_{26}(\mathbf{X}_t) = \gamma I_2, \\
&\mathcal{E}_{27} : A_2^* \to R_2^*, \qquad h_{27}(\mathbf{X}_t) = \rho A_2^*, \\
&\mathcal{E}_{28} : A_2^* \to D_2^*, \qquad h_{28}(\mathbf{X}_t) = \delta_v^2 \delta A_2^*.
\end{aligned}
$$

The respective state change vectors $\boldsymbol{\nu}_1, \boldsymbol{\nu}_2, \ldots, \boldsymbol{\nu}_{28}$ can be obtained in a straightforward manner based on the ordering of compartments in the full system state vector is $\mathbf{X}_t =$

$[S_{u,t}, E_{u,t}, I_{u,t}, R_{u,t}, A^*_{u,t}, R^*_{u,t}, D^*_{u,t}, S_{1,t}, \ldots, D^*_{1,t},\ S_{2,t}, \ldots, D^*_{2,t}]$. The observable data are $\mathbf{Z}_t = [C^*_t, D^*_t, V^*_{1,t}, V^*_{2,t}]$ with

$$
\begin{aligned}
C^*_t &= A^*_{u,t} + A^*_{1,t} + A^*_{2,t} + R^*_{u,t} + R^*_{1,t} + R^*_{2,t} \\
D^*_t &= D^*_{u,t} + D^*_{1,t} + D^*_{2,t} \\
V^*_{1,t} &= S_{1,t} + E_{1,t} + I_{1,t} + R_{1,t} + A^*_{1,t} + R^*_{1,t} + D^*_{1,t} \\
V^*_{2,t} &= S_{2,t} + E_{2,t} + I_{2,t} + R_{2,t} + A^*_{2,t} + R^*_{2,t} + D^*_{2,t}.
\end{aligned}
$$

The response function $g(\mathbf{Z}_t, t)$ and hesitancy effect function $h(\mathbf{Z}_t, t)$ are given by,

$$
\begin{aligned}
g(\mathbf{Z}_t, t) &= \frac{1}{1 + (\mathbb{1}_{[0,T_d]}(t)\, w_A \zeta C^*_t)^n}, \\
h(\mathbf{Z}_t, t) &= \frac{(w_C C^* + w_D D^* + w_V V^*_2)^{n_v}}{1 + (w_C C^* + w_D D^* + w_V V^*_2)^{n_v}} \mathbb{1}_{[T_v,\infty)}(t)\,.
\end{aligned}
$$

These correspond to Equations (2) and (6) in the main manuscript.

#### A.3 Stochastic Simulation

While exact realisations of this process can be generated using event-based simulation [3, 4], this is prohibitive within an approximate Bayesian computational setting with large population sizes and event numbers. Therefore, we apply a first order approximation to the integral over the interval $[t, t+\tau)$ to obtain the tau-leaping approximation [5],

$$
\mathbf{X}_{t+\tau} = \mathbf{X}_t + \sum_{j=1}^{28} Y_j \boldsymbol{\nu}_j + \mathcal{O}(\tau),
$$

where $Y_j \sim \text{Poisson}\,(h_j(\mathbf{X}_t)\tau)$ counts the number of times event $j$ occurs in the interval $[t, t+\tau)$. Simulations proceed as per Gillespie [5]. For our simulations we use $\tau = 1$ (days), and initial condition $\mathbf{X}_0 = [P - (3\kappa\zeta + 1)C^*_0 - D^*_0, 2\kappa\zeta C^*_0, \kappa C^*_0, 0, \zeta C^*_0, (1-\zeta)C^*_0, D^*_0, \mathbf{0}]^{\mathrm{T}}$.

### Appendix B Approximate Bayesian analysis

We apply Bayesian inference to quantify uncertainty in the model parameters associated vaccine hesitancy effects, $\boldsymbol{\theta} = [\nu, n_n, w_C, w_D, w_V]$, for each synthetic dataset $i$ that represents the structure of data sets available from Johns Hopkins University or Our World in Data dashboards, that is, $\mathcal{D}_i = [\{C^*_{t,i}, D^*_{t,i}, V^*_{1,t,i}, V^*_{2,t,i}\}_{T \geq t \geq 0}]$ where $T$ is the total number of days available of data. The task is to sample the posterior distribution with probability density given by Bayes' Theorem,

$$p(\boldsymbol{\theta} \mid \mathcal{D}_i) = \frac{p(\mathcal{D}_i \mid \boldsymbol{\theta})p(\boldsymbol{\theta})}{p(\mathcal{D}_i)},$$

where $p(\boldsymbol{\theta})$ is the prior, $p(\mathcal{D}_i \mid \boldsymbol{\theta})$ is the likelihood and $p(\mathcal{D}_i)$ is the evidence. For the remainder of this section, we omit the country index $i$ for notational convenience.

Since the full model state vector is only partially observable, the data model is no longer Markovian and is therefore computationally intractable. To deal with this likelihood intractability, we apply approximate Bayesian computation (ABC) [7–9], that samples from an approximation to the posterior for each country,

$$\begin{aligned} p(\boldsymbol{\theta} \mid \mathcal{D}) \approx p(\boldsymbol{\theta} \mid \rho(\mathcal{D}, \mathcal{D}_s) \leq \epsilon) \propto \mathbb{P}(\rho(\mathcal{D}, \mathcal{D}_s) \leq \epsilon \mid \boldsymbol{\theta})p(\boldsymbol{\theta}) \\ = p(\boldsymbol{\theta}) \int \mathbb{1}_{(0,\epsilon]}\left(\rho(\mathcal{D}, \mathcal{D}_s)\right) s(\mathcal{D}_s \mid \boldsymbol{\theta}) \, \mathrm{d}\mathcal{D}_s, \end{aligned}$$

where $\mathcal{D}$ is the synthetic data that will align to one of the hesitancy scenarios scenario, $\mathcal{D}_s \sim s(\cdot \mid \boldsymbol{\theta})$ is simulated data from the full model, $\rho(\mathcal{D}, \mathcal{D}_s)$ is a discrepancy metric, $\epsilon$ is the discrepancy threshold and $\mathbb{1}_{(0,\epsilon]}\left(\rho(\mathcal{D}, \mathcal{D}_s)\right) = 1$ if $\rho(\mathcal{D}, \mathcal{D}_s) \leq \epsilon$, and $\mathbb{1}_{(0,\epsilon]}\left(\rho(\mathcal{D}, \mathcal{D}_s)\right) = 0$ otherwise. For our implementation, we apply the discrepancy metric,

$$\rho(\mathcal{D}, \mathcal{D}_s) = \left( \sum_{t=1}^{T} (C^*_t - C^*_{t,s})^2 + (D^*_t - D^*_{t,s})^2 + (V^*_{1,t} - V^*_{1,t,s})^2 + (V^*_{2,t} - V^*_{2,t,s})^2 ) \right)^{1/2}$$

where $\mathcal{D} = [\{A^*_t, D^*_t, V^*_{1,t}, V^*_{2,t}\}_{t \geq 0}]$ is the synthetic data for a given scenario and $\mathcal{D}_s = [\{A^*_{t,s}, D^*_{t,s}, V^*_{1,t,s}, V^*_{2,t,s}\}_{t \geq 0}]$ is simulated data form the full model.

#### B.1 Sequential Monte Carlo sampling

We apply a sequential Monte Carlo (SMC) scheme [1,6] to move an initial set of $N_p$ samples from the prior through a sequence of ABC approximations defined by a decreasing sequence of $T$ discrepancy thresholds, $\epsilon_1 > \epsilon_2 > \cdots > \epsilon_T = \epsilon$. Our particular implementation (Algorithm 1), based on the work of Drovandi and Pettit [2], adaptively selects the acceptance thresholds and utilises MCMC steps using tuned Gaussian random walk proposals. For all model calibrations we apply adaptive SMC with $N_p = 1000$ particles, tuning parameters $c = 0.01$, $a = 0.5$ and terminate sampling when the MCMC acceptance probability $p_{\text{acc}}$ drops below $p_{\min} = 0.01$. The acceptance probability is estimated using $R_{\text{trial}} = 5$ initial MCMC iterations per particle.

**Algorithm 1** Adaptive SMC sampler for approxmate Bayesian computation

Initialise $N_a = aN_p$, $N_\ell = N_p - N_a$
**for** $j \in [1, 2, \dots, N_p]$ **do**
    Sample prior, $\boldsymbol{\theta}^* \sim p(\cdot)$ and simulate model, $\mathcal{D}_s \sim s(\cdot \mid \boldsymbol{\theta}^*)$;
    Set $\rho_j \leftarrow \rho(\mathcal{D}, \mathcal{D}_s)$;
**end for**
**repeat**
    Sort particles $\{(\boldsymbol{\theta}_j, \rho_j)\}_{j=1}^{N_p}$, such that $\rho_j \leq \rho_{j+1}$ for all $j \in [1, 2, \dots, N_p - 1]$;
    Remove particles $\{(\boldsymbol{\theta}_j, \rho_j)\}_{j=N_\ell+1}^{N_p}$ an set $\epsilon \leftarrow \rho_{N_\ell}$;
    Resample particles $\{\boldsymbol{\theta}_j\}_{j=N_\ell+1}^{N_p}$ from $\{(\boldsymbol{\theta}_j)\}_{j=1}^{N_\ell}$ with replacement;
    Estimate sample covariance, $\hat{\boldsymbol{\Sigma}}$, of particles $\{\boldsymbol{\theta}_j\}_{j=1}^{N_p}$.
    Adapt proposal kernel $q(\mathbf{u} \mid \mathbf{v}) = \phi\left(\mathbf{u}; \mathbf{v}, \dfrac{2.38^2}{\dim(\boldsymbol{\theta})}\hat{\boldsymbol{\Sigma}}\right)$, where $\phi(\cdot; \boldsymbol{\mu}, \boldsymbol{\Sigma})$ is a multivariate Gaussian density function and $\dim(\boldsymbol{\theta})$ is the number of parameters;
    Set $p_{\text{acc}} \leftarrow 0$;
    **for** $j \in [N_\ell + 1, N_\ell + 2, \dots, N_p]$ **do**
        **for** $k \in [1, 2, \dots, R_{\text{trial}}]$ **do**
            Generate proposal, $\boldsymbol{\theta}^* \sim q(\cdot \mid \boldsymbol{\theta}_j)$ and sample $u \sim \mathcal{U}(0, 1)$;
            **if** $u \leq \min\left(1, \dfrac{p(\boldsymbol{\theta}^*)q(\boldsymbol{\theta}_j \mid \boldsymbol{\theta}^*)}{p(\boldsymbol{\theta}_j)q(\boldsymbol{\theta}^* \mid \boldsymbol{\theta}_j)}\right)$ **then**
                Simulate model $\mathcal{D}_s \sim s(\cdot \mid \boldsymbol{\theta}^*)$;
                **if** $\rho(\mathcal{D}_i, \mathcal{D}_s) \leq \epsilon$ **then**
                    Set $\boldsymbol{\theta}_j \leftarrow \boldsymbol{\theta}^*$, $\rho_j \leftarrow \rho(\mathcal{D}_i, \mathcal{D}_s)$, and $p_{\text{acc}} \leftarrow p_{\text{acc}} + (R_{trial} N_a)^{-1}$;
                **end if**
            **end if**
        **end for**
    **end for**
    Set $R \leftarrow \log c / \log(1 - p_{\text{acc}})$;
    **for** $j \in [N_\ell + 1, N_\ell + 2, \dots, N_p]$ **do**
        **for** $k \in [1, 2, \dots, R - R_{\text{trial}}]$ **do**
            Generate proposal, $\boldsymbol{\theta}^* \sim q(\cdot \mid \boldsymbol{\theta}_j)$ and sample $u \sim \mathcal{U}(0, 1)$;
            **if** $u \leq \min\left(1, \dfrac{p(\boldsymbol{\theta}^*)q(\boldsymbol{\theta}_j \mid \boldsymbol{\theta}^*)}{p(\boldsymbol{\theta}_j)q(\boldsymbol{\theta}^* \mid \boldsymbol{\theta}_j)}\right)$ **then**
                Simulate model $\mathcal{D}_s \sim s(\cdot \mid \boldsymbol{\theta}^*)$;
                **if** $\rho(\mathcal{D}_i, \mathcal{D}_s) \leq \epsilon$ **then**
                    Set $\boldsymbol{\theta}_j \leftarrow \boldsymbol{\theta}^*$, $\rho_j \leftarrow \rho(\mathcal{D}_i, \mathcal{D}_s)$, and $p_{\text{acc}} \leftarrow p_{\text{acc}} + (R N_a)^{-1}$;
                **end if**
            **end if**
        **end for**
    **end for**
**until** $p_{\text{acc}} < p_{\min}$

### Appendix C Additional Results

Figures 1–8 show equivalent Bayesian analyses results for other synthetic data set configurations. The patterns related to bivariate marginal posterior correlation structures are consistent with those discussed in the main manuscript.

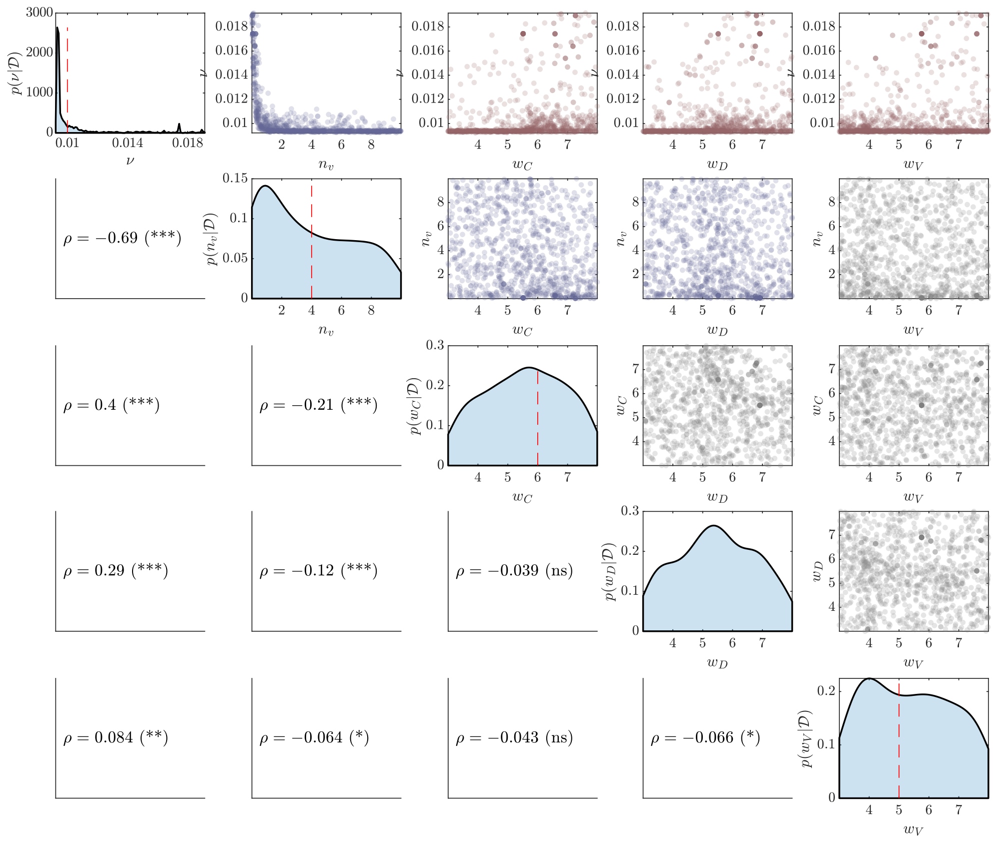

Figure 1: Example posterior samples for the simulation scenario with $\nu = 0.01$, $n_v = 4$, $w_C = 10^{-6}$, $w_D = 0$ and $w_V = 2 \times 10^{-6}$. The main diagonals show marginal posterior densities with dashed red lines indicating the true values. Marginals without a dashed red line indicates the true parameter corresponds to $w_C, w_D, w_V \to \infty$ or $w_C, w_D, w_V \to 0$. The upper off-diagonal plots show the bivariate posterior samples, from which the pairwise correlations are computed. Plots with significantly positive, significantly negative and statistically insignificant correlations are shown in red, blue and grey respectively. The lower off-diagonal plots show the Spearman rank correlation and statistical significance level.

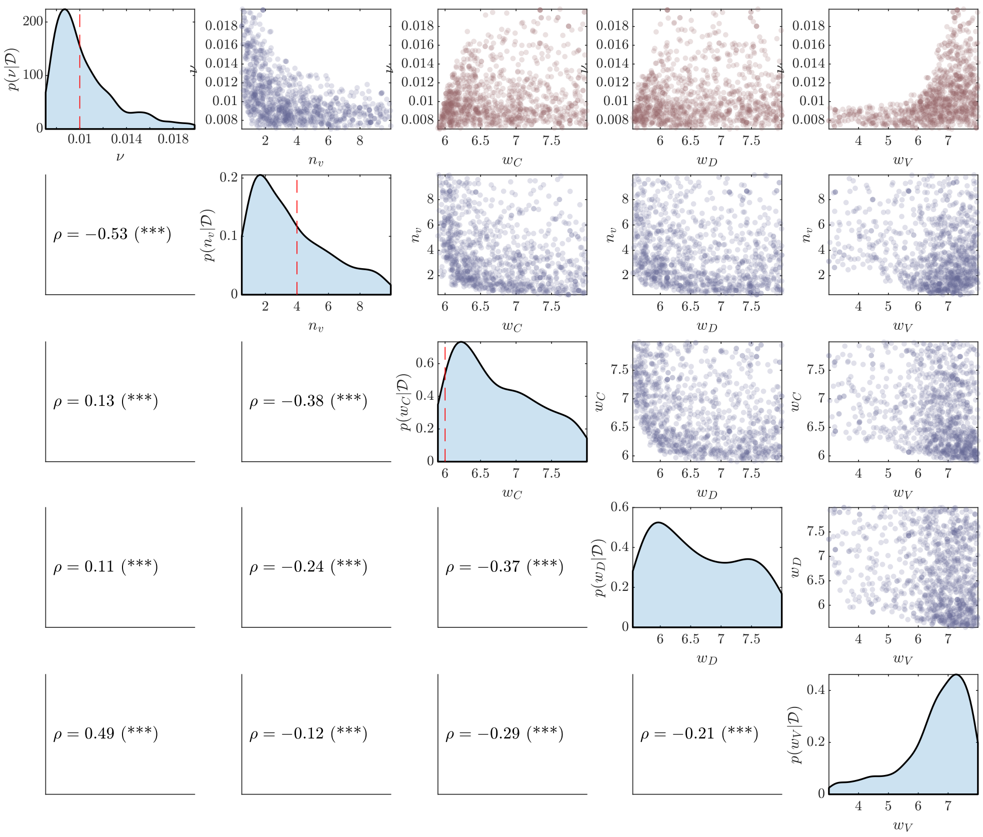

Figure 2: Example posterior samples for the simulation scenario with $\nu = 0.01$, $n_v = 4$, $w_C = 10^{-6}$, $w_D = 0$ and $w_V = 0$. The main diagonals show marginal posterior densities with dashed red lines indicating the true values. Marginals without a dashed red line indicates the true parameter corresponds to $w_C, w_D, w_V \to \infty$ or $w_C, w_D, w_V \to 0$. The upper off-diagonal plots show the bivariate posterior samples, from which the pairwise correlations are computed. Plots with significantly positive, significantly negative and statistically insignificant correlations are shown in red, blue and grey respectively. The lower off-diagonal plots show the Spearman rank correlation and statistical significance level.

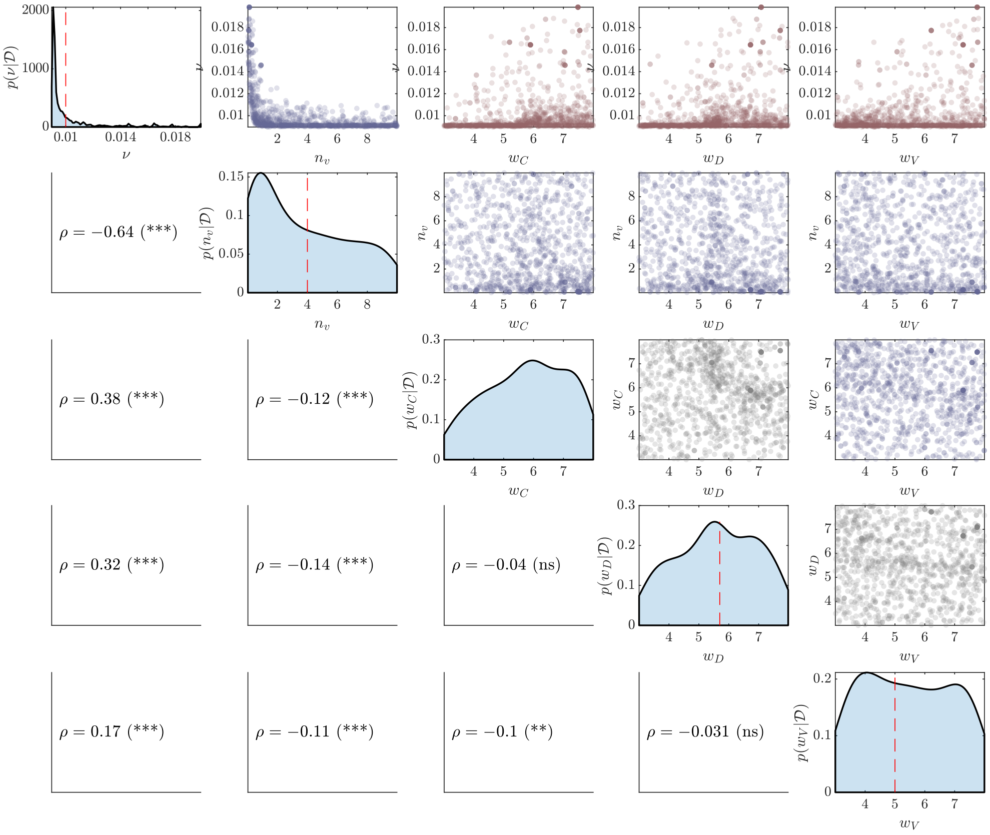

Figure 3: Example posterior samples for the simulation scenario with $\nu = 0.01$, $n_v = 4$, $w_C = 0$, $w_D = 2 \times 10^{-6}$ and $w_V = \times 10^{-5}$. The main diagonals show marginal posterior densities with dashed red lines indicating the true values. Marginals without a dashed red line indicates the true parameter corresponds to $w_C, w_D, w_V \to \infty$ or $w_C, w_D, w_V \to 0$. The upper off-diagonal plots show the bivariate posterior samples, from which the pairwise correlations are computed. Plots with significantly positive, significantly negative and statistically insignificant correlations are shown in red, blue and grey respectively. The lower off-diagonal plots show the Spearman rank correlation and statistical significance level.

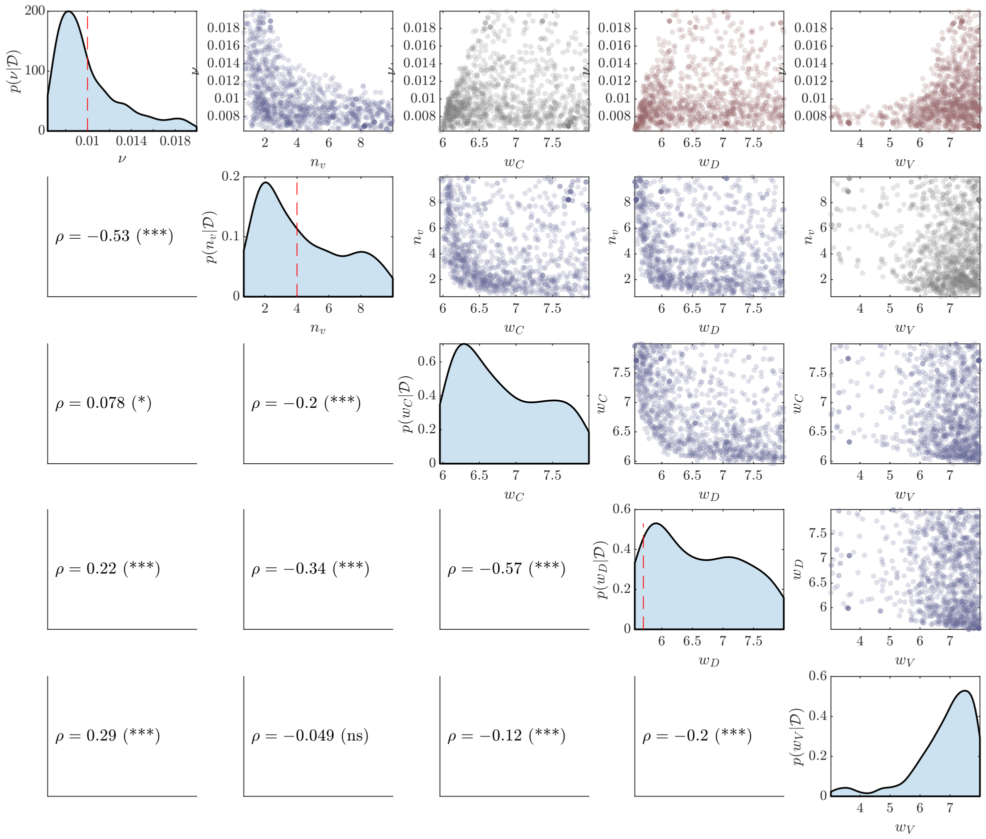

Figure 4: Example posterior samples for the simulation scenario with $\nu = 0.01$, $n_v = 4$, $w_C = 0$, $w_D = 2\times10^{-6}$ and $w_V = 0$. The main diagonals show marginal posterior densities with dashed red lines indicating the true values. Marginals without a dashed red line indicates the true parameter corresponds to $w_C, w_D, w_V \to \infty$ or $w_C, w_D, w_V \to 0$. The upper off-diagonal plots show the bivariate posterior samples, from which the pairwise correlations are computed. Plots with significantly positive, significantly negative and statistically insignificant correlations are shown in red, blue and grey respectively. The lower off-diagonal plots show the Spearman rank correlation and statistical significance level.

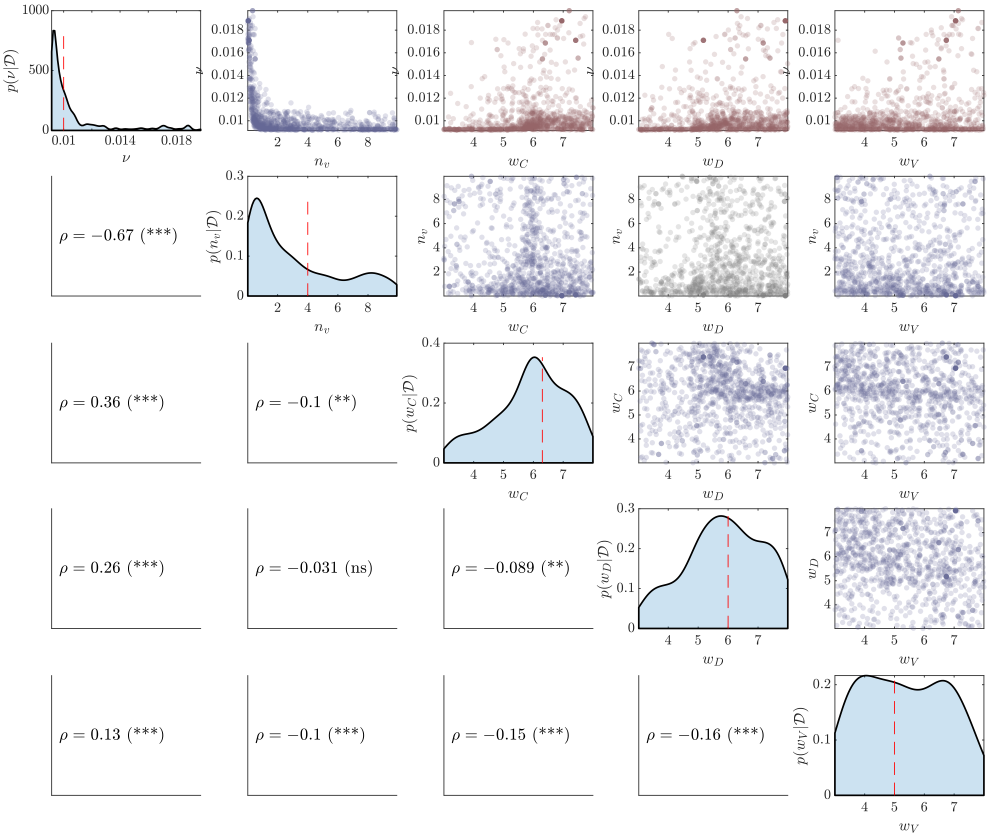

Figure 5: Example posterior samples for the simulation scenario with $\nu = 0.01$, $n_v = 4$, $w_C = 5\times10^{-7}$, $w_D = \times10^{-6}$ and $w_V = 10^{-5}$. The main diagonals show marginal posterior densities with dashed red lines indicating the true values. Marginals without a dashed red line indicates the true parameter corresponds to $w_C, w_D, w_V \to \infty$ or $w_C, w_D, w_V \to 0$. The upper off-diagonal plots show the bivariate posterior samples, from which the pairwise correlations are computed. Plots with significantly positive, significantly negative and statistically insignificant correlations are shown in red, blue and grey respectively. The lower off-diagonal plots show the Spearman rank correlation and statistical significance level.

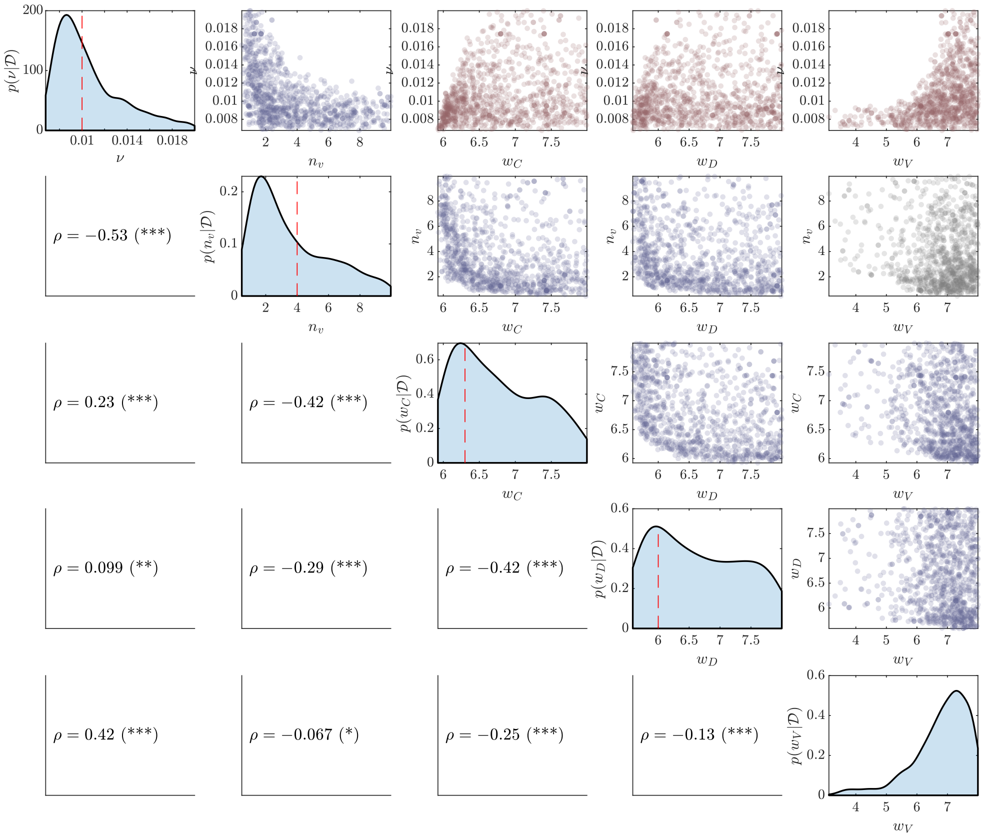

Figure 6: Example posterior samples for the simulation scenario with $\nu = 0.01$, $n_v = 4$, $w_C = 5 \times 10^{-7}$, $w_D = \times 10^{-6}$ and $w_V = 0$. The main diagonals show marginal posterior densities with dashed red lines indicating the true values. Marginals without a dashed red line indicates the true parameter corresponds to $w_C, w_D, w_V \to \infty$ or $w_C, w_D, w_V \to 0$. The upper off-diagonal plots show the bivariate posterior samples, from which the pairwise correlations are computed. Plots with significantly positive, significantly negative and statistically insignificant correlations are shown in red, blue and grey respectively. The lower off-diagonal plots show the Spearman rank correlation and statistical significance level.

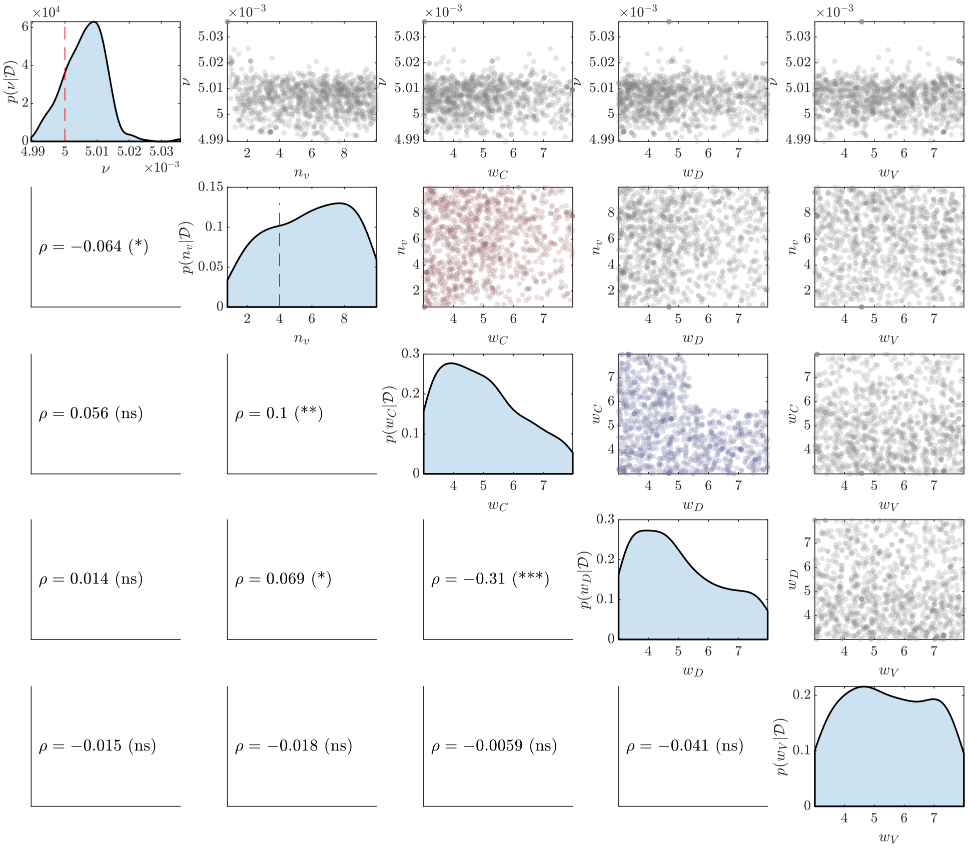

Figure 7: Example posterior samples for the simulation scenario with $\nu = 0.005$, $n_v = 4$, $w_C to\infty$, $w_D = 0$ and $w_V = 0$. The main diagonals show marginal posterior densities with dashed red lines indicating the true values. Marginals without a dashed red line indicates the true parameter corresponds to $w_C, w_D, w_V \to \infty$ or $w_C, w_D, w_V \to 0$. The upper off-diagonal plots show the bivariate posterior samples, from which the pairwise correlations are computed. Plots with significantly positive, significantly negative and statistically insignificant correlations are shown in red, blue and grey respectively. The lower off-diagonal plots show the Spearman rank correlation and statistical significance level.

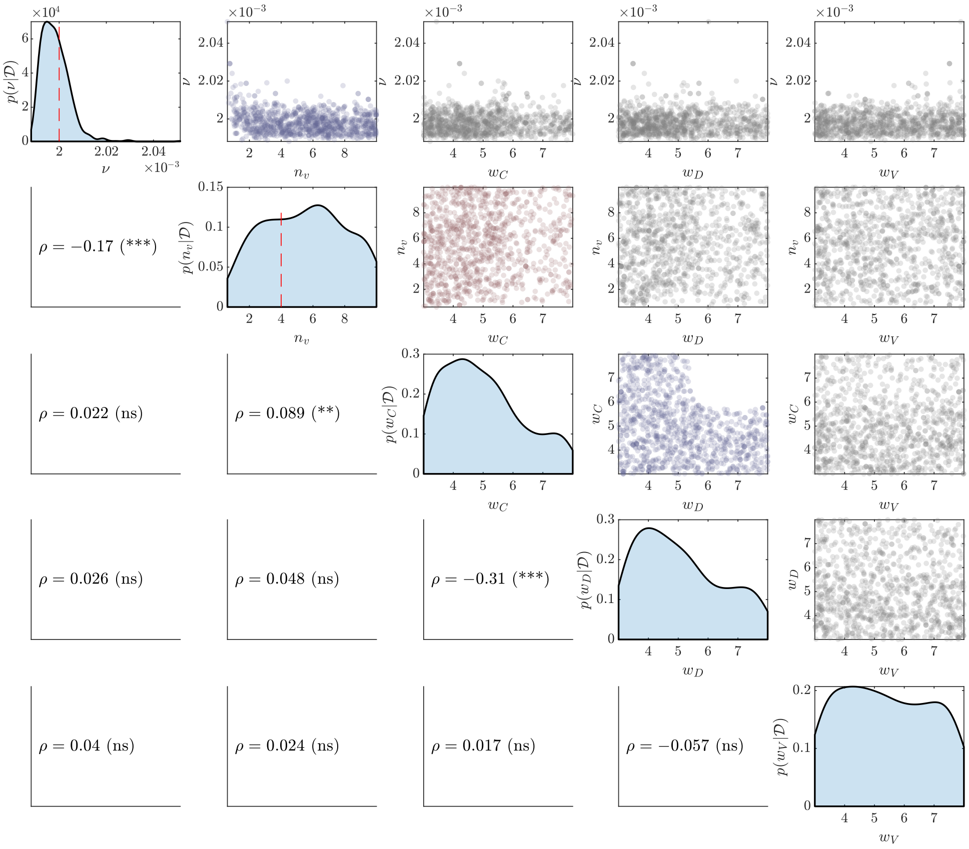

Figure 8: Example posterior samples for the simulation scenario with $\nu = 0.002$, $n_v = 4$, $w_C to \infty$, $w_D = 0$ and $w_V = 0$. The main diagonals show marginal posterior densities with dashed red lines indicating the true values. Marginals without a dashed red line indicates the true parameter corresponds to $w_C, w_D, w_V \to \infty$ or $w_C, w_D, w_V \to 0$. The upper off-diagonal plots show the bivariate posterior samples, from which the pairwise correlations are computed. Plots with significantly positive, significantly negative and statistically insignificant correlations are shown in red, blue and grey respectively. The lower off-diagonal plots show the Spearman rank correlation and statistical significance level.
